# Outcome Reporting bias in Exercise Oncology trials (OREO): a cross-sectional study

**DOI:** 10.1101/2021.03.12.21253378

**Authors:** Benjamin Singh, Ciaran M Fairman, Jesper F Christensen, Kate A Bolam, Rosie Twomey, David Nunan, Ian M Lahart

**Affiliations:** Greenslopes Private Hospital, Brisbane, Australia; Exercise Science, Arnold School of Public Health, University of South Carolina, South Carolina, USA; Centre for Physical Activity Research, Rigshospitalet, Copenhagen, Denmark; Department of Sports Science and Clinical Biomechanics, Faculty of Health Sciences, University of Southern Denmark, Denmark; Department of Neurobiology, Care Sciences and Society, Karolinska Institutet, Stockholm, Sweden; Cumming School of Medicine, University of Calgary, Calgary, Canada; Nuffield Department of Primary Care Health Sciences, University of Oxford, Oxford, UK; Sport and Physical Activity Research Centre, Faculty of Education, Health, and Wellbeing, University of Wolverhampton, Wolverhampton, UK

**Author notes:** Corresponding author: Ian M Lahart.

## Abstract

**Background:** Despite evidence of selective outcome reporting across multiple disciplines, this has not yet been assessed in trials studying the effects of exercise in people with cancer. Therefore, the purpose of our study was to explore prospectively registered randomised controlled trials (RCTs) in exercise oncology for evidence of selective outcome reporting.

**Methods:** Eligible trials were RCTs that 1) investigated the effects of at least partially supervised exercise interventions in people with cancer; 2) were preregistered (i.e. registered before the first patient was recruited) on a clinical trials registry; and 3) reported results in a peer-reviewed published manuscript. We searched the PubMed database from the year of inception to September 2020 to identify eligible exercise oncology RCTs clinical trial registries. Eligible trial registrations and linked published manuscripts were compared to identify the proportion of sufficiently preregistered outcomes reported correctly in the manuscripts, and cases of outcome omission, switching, and silently introduction of non-novel outcomes.

**Results:** We identified 31 eligible RCTs and 46 that were ineligible due to retrospective registration. Of the 405 total prespecified outcomes across the 31 eligible trials, only 6.2% were preregistered complete methodological detail. Only 16% (n=148/929) of outcomes reported in published results manuscripts were linked with sufficiently preregistered outcomes without outcome switching. We found 85 total cases of outcome switching. A high proportion (41%) of preregistered outcomes were omitted from the published results manuscripts, and many published outcomes (n=394; 42.4%) were novel outcomes that had been silently introduced (median, min-max=10, 0-50 per trial). We found no examples of preregistered efficacy outcomes that were measured, assessed, and analysed as planned.

**Conclusions:** We found evidence suggestive of widespread selective outcome reporting and non-reporting bias (outcome switching, omitted preregistered outcomes, and silently introduced novel outcomes). The existence of such reporting discrepancies has implications for the integrity and credibility of RCTs in exercise oncology.

**Preregistered protocol:** https://osf.io/dtkar/ (posted: November 19, 2019)

---

> *“OF COOKING. This is an art of various forms, the object of which is to give to ordinary observations the appearance and character of those of the highest degree of accuracy. One of its numerous processes is to make multitudes of observations, and out of these to select those only which agree, or very nearly agree. If a hundred observations are made, the cook must be very unlucky if he cannot pick out fifteen or twenty which will do for serving up.”*

Charles Babbage, Reflections on the decline of science in England, and on some of its causes, 29^th^ April 1830 (1).

## INTRODUCTION

Exercise oncology—the study of exercise in people living with and beyond cancer—has witnessed an exponential growth in research over the last 30 years. Hundreds of randomized controlled trials (RCTs) examining the feasibility, safety, efficacy, or effectiveness of exercise interventions before, during, and after active oncology treatment across different cancer types have contributed to an ever-growing multitude of systematic reviews and meta-analyses, which in turn have been synthesized in umbrella reviews (2, 3). Such evidence now underpins clinical exercise guideline recommendations, including specific exercise prescriptions for some cancer-related health outcomes (4, 5) and appeals for exercise to be viewed as standard practice in oncology (6, 7).

RCTs are considered the optimal design to assess whether health interventions offer a benefit or cause harm, and therefore, provide important evidence for clinical decision-making (8). However, reporting biases such as selective (non-) reporting of outcomes (see Box 1 for definitions) threaten the validity of RCTs, and therefore the reviews and guidelines they inform. The prevalence and impact of these outcome reporting biases in the wider published biomedical literature has been well studied and highlights an abundance of potential false positive findings, interventions with likely overestimated benefits and underestimated harms, and meta-analyses overwhelmingly in favour of studied interventions (9–14).

#### Box 1. Definitions of reporting biases [definitions taken from: (13, 15)]

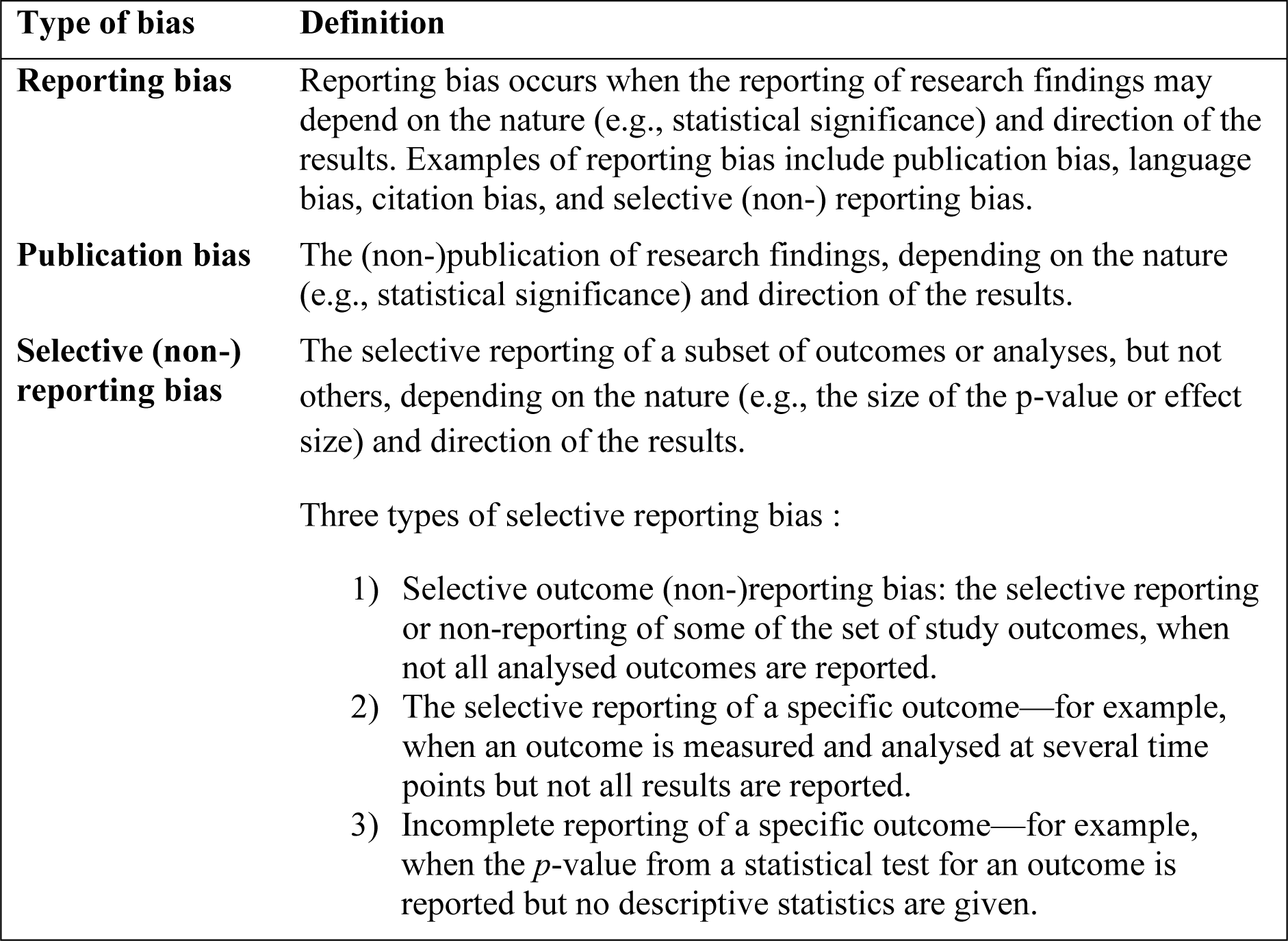

Although selective reporting bias, or ‘cooking’ as Charles Babbage dubbed it in 1830, is a long-standing problem in science publication, direct evidence of its existence has only surfaced more recently. In a landmark paper in 2004, Chan and colleagues (10) discovered that 62% of RCTs approved by the Scientific-Ethical Committees for Copenhagen and Frederiksberg, Denmark, had at least one primary outcome that was switched for a non-secondary outcome (i.e. outcome switching), silently introduced, or omitted when they compared published trial manuscripts with their accompanying protocols. The authors (10) concluded that to “ensure transparency, planned trials should be registered and protocols should be made publicly available prior to trial completion” (pp. 2457).

Several initiatives now exist to explicitly address the negative impacts of selective reporting bias. For over a decade, an international standard for clinical trials is prospective registration in a public database before recruitment of the first participant (16–18). This is mandated by the Declaration of Helsinki (17), endorsed by the World Health Organization (WHO) (19), and a requirement for publication of trial findings in member journals of the International Committee of Medical Journal Editors (ICMJE) (18). Clinical trial registration aims to track all initiated trials, identify unpublished trials, achieve transparency in trial reporting (a fundamental tenet of science), and make key information about the objectives and design of a trial publicly available (12,20,21). A prospective registration also allows for the distinction between confirmatory outcomes and those which are exploratory (22, 23). Although trial registries have served as an audit trail for documenting discrepancies between registrations and trial publications, they do not appear to have acted as a deterrent to selective reporting biases (12,24–26).

In 2019, Goldacre and colleagues (26) provided strong evidence that selective outcome reporting is still a major concern even in those trials published in so-called prestige journals. The authors (26) prospectively assessed outcome reporting in all newly published trials in five of the highest impact medical journals over a 6-week period, and revealed that 87% of trials had discrepancies serious enough to warrant a correction letter, and five undeclared outcomes were on average added per trial. Similar evidence of reporting discrepancies has been observed across many disciplines (24), including psychology (25), cognitive science (27), cystic fibrosis (28), anaesthesiology (29), drug trials (30, 31), and oncology (32, 33).

Exercise oncology shares with these disciplines many of the features that can propagate selective (non-) reporting bias, such as a preponderance of studies with positive or favourable results, non-cooperative group trials, assessment of a vast array of outcomes from a multitude of measurement tools, large researcher degrees of freedom when choosing between outcome and analysis options, and high pressures to publish (27,34–36). Given these similarities, it seems unreasonable to believe that exercise oncology research is immune to the problems that have contributed to a ‘credibility crisis’ in other disciplines (37).

A recent umbrella review reported that the effect size estimates are typically small-moderate, and the certainty of the evidence is typically graded as low or moderate across meta-analyses of exercise oncology RCTs (3). Consequently, many effects within these meta-analyses may be vulnerable to selective (non-) reporting bias, which could change the direction, size, and certainty of pooled effect estimates. The existence of selective (non-) reporting in exercise oncology trials risks undermining exercise-based clinical guidelines and calls to include exercise in the standard care of patients with cancer (6). To date, however, selective (non-) reporting biases have not been assessed in RCTs of exercise interventions for people diagnosed with cancer. Therefore, the overall purpose of this study is to evaluate selective (non-) reporting in prospectively registered RCTs in the field of exercise oncology.

## METHOD

Our study protocol was preregistered on November 19, 2019 and can be accessed here: https://osf.io/dtkar/.

### Search and trial eligibility

Eligible trials: 1) were longitudinal RCTs investigating the efficacy or effectiveness of at least partially supervised exercise interventions on health-related outcomes in people diagnosed with cancer; 2) were preregistered on either ClinicalTrials.gov, the International Standard Randomised Controlled Trials Number (ISRCTN) registry via the World Health Organization International Clinical Trials Registry Platform (WHO ICTRP), the European Union (EU) Clinical Trials Register, the Australian and New Zealand Clinical Trials Registry (ANZCTR), Netherlands Trial Registry (NTR), the Chinese Clinical Trial Registry (ChiCTR), or the UMIN Clinical Trials Registry (UMIN-CTR); and 3) reported their findings in at least one full-text article in a peer-reviewed journal. We did not exclude trials based on cancer type or stage, or treatment status (e.g., during or following adjuvant therapy, pre- or post-surgery).

In line with the ICMJE’s definition of a clinical trial (18), we defined an exercise oncology RCT as a research project that prospectively and randomly assigned individuals diagnosed with cancer to intervention or comparison groups to study the cause-and-effect relationship between an exercise intervention and a health outcome. We adopted the following definition of exercise: “a potential disruption to homeostasis by muscle activity that is either exclusively, or in combination, concentric, eccentric or isometric muscle contractions” (38). We included both efficacy studies that examined the benefits or harms of an exercise intervention under controlled conditions and effectiveness or pragmatic trials that investigated exercise interventions under conditions that were closer to real-world practice (39), if they involved some level of face-to-face supervision. We excluded behaviour-based trials (i.e. studies that focused primarily on increasing physical activity). Exercise interventions that involved a behaviour change component, however, were included if some level of face-to-face exercise supervision was involved. Trials that evaluated drug, dietary, supplement, or alternative interventions in combination with exercise were eligible only if the effects of exercise could be isolated (i.e. if a comparison group also receives the drug or supplement without exercise; e.g., an exercise plus drug arm *vs.* a drug only arm).

We defined a preregistered trial as a trial that was prospectively registered (i.e. the trial was registered on or before the stated study start date or onset of patient enrolment) on any of the clinical trial registries listed above (we accepted trials that had a start date the same month as they were registered, if the day of the month was not reported) (40). We excluded, but made note of, retrospectively registered trials (i.e. those that were registered during or after the completion of the study), because they may have been registered after an interim or final data analyses, allowing for selective registration of certain outcomes dependent on the outcome direction or statistical significance. Additionally, papers linked to eligible trials that reported only the results of cross-sectional analyses were excluded if they did not compare the effects of an eligible exercise intervention on outcomes versus comparison groups.

## Outcomes

Primary outcome:

The proportion of sufficiently preregistered primary and secondary outcomes reported correctly in published manuscripts.

- Outcome preregistration was considered ‘sufficiently’ reported when the preregistered outcome indicated the outcome score (e.g., the FACT-general total score is an outcome score used as a measure of quality of life) and not only the outcome domain (e.g., registering “quality of life” alone was considered insufficient).
- ‘Correctly’ reported outcomes were outcomes that were preregistered primary outcomes reported in publications as a primary outcome or preregistered secondary outcomes reported in publications as secondary outcomes, or in cases where the authors declared and provided a justification for discrepancies between the preregistration and published manuscript.

Secondary outcomes:

1. The proportion of (both sufficiently preregistered and domain only) primary outcomes omitted or reported as a secondary outcome in the published manuscripts (i.e. outcome switching).
2. The proportion of (both sufficiently preregistered and domain only) secondary outcomes omitted or reported as primary outcomes in the published manuscripts (i.e. outcome switching).
3. The total number of instances of outcome switching in preregistered primary and secondary outcomes.
4. The number of undeclared and non-preregistered (novel) outcomes added to published manuscripts (i.e. silently introduced).
5. The proportion of sufficiently preregistered outcomes with descriptions in published manuscripts consistent with their preregistered descriptions (i.e. method of assessment, outcome score, assessment timepoints, and statistical analysis plan matches the description in the published manuscripts).

### Search of trials

We undertook a PubMed database search to identify all potentially eligible exercise RCTs involving patients diagnosed with cancer using registry-specific search terms (Secondary Source IDs) for the following registries: ClinicalTrials.gov, ISRCTN registry, the WHO ICTRP, the EU Clinical Trials Register, ANZCTR, NTR, ChiCTR, and the UMIN-CTR. Details of the PubMed search strategy are presented in Supplementary Table 1. We searched for trials registered from the year of database inception to November 2019 (and updated September 2020). Of note, the earliest trial registry started in 2000 [ClinicalTrials.gov, (41)], and registries were seldom used prior to the 2004 ICMJE statement requiring trial registration (40).

One investigator (IML) undertook the registry searches and uploaded all records identified during the search to Rayyan systematic review software [Qatar Computing Research Institute (Data Analytics)]. We searched the WHO ICTRP registry (no Secondary Source ID was available for this registry) separately using a title and keyword search (Supplementary Table 1) and exported the search results into Microsoft Excel™. Two investigators (BS and CMF) independently screened all records detected during the registry search (in Rayyan and Microsoft Excel) to identify potentially eligible trials applying the eligibility criteria described above. A third investigator (IML) resolved any discrepancies.

We identified published manuscripts linked to each trial via a PubMed and Google Scholar search (November 2019, and updated in September 2020) using the trial registry code (e.g., NCT number), and a search of publication lists on the trial registrations (e.g., Clinicaltrials.gov lists publications automatically indexed to a study by NCT Number). We also searched for a prospectively published version of a protocol for each trial that was dated before the trial start date (i.e. preregistered). Where a prospective trial registration was accompanied by a preregistered published protocol paper, the trial registration was used an *a priori*. If the protocol did not sufficiently define the prespecified outcomes, the registry entry was used instead, and vice versa [for only three trials, (42–44)], additional information was taken from the published protocol due to a lack of detail in the registry entries). We also accessed supplementary material linked to a published article to extract the required data.

### Data extraction and coding

Five investigators (BS, CMF, IML, DN, and KAB) extracted all relevant information from the eligible studies and recorded this on specifically designed data forms (https://bit.ly/3qFkcro). From the registration entry of each trial, the following information was extracted: registration date, trial start date, current status, onset of participant enrolment, primary completion date (i.e. date of final collection of data for the primary outcome), nature and number of the reported outcome measures with their methods and time frame for data collection, timepoints at which the outcomes are assessed, planned statistical analysis (including power analysis), and registered changes in outcome measures (or trial procedures) with corresponding dates. Next, across all eligible publications and associated supplementary materials, we (BS, CMF, IML, and KAB) extracted patient, cancer (e.g., sample size details, patients’ gender, cancer type, and treatment status), trial, and intervention characteristics (trial duration, condition numbers and types, and exercise type), matched the reporting of outcomes prespecified in the registries to that of the publications, and noted the *p*-values, effect sizes, and descriptive statistics [e.g., mean differences between groups with 95% confidence intervals (CIs)] of all outcomes reported.

### Assessing the completeness of preregistrations

We assessed the completeness of preregistrations using a modified version of Zarin et al.’s (45) “four levels of specification in reporting outcome measures” method. Specifically, we established whether authors preregistered the following: 1) outcome domain (i.e. broad categories of outcomes that can be represented by multiple outcome scores measured via different instruments or scales, e.g., quality of life); 2) outcome measure method (e.g., FACT-General questionnaire); 3) outcome assessment timepoints (e.g., baseline and post-intervention at 12 weeks); 4) outcome score (i.e. the score that would be entered into a subsequent statistical analysis, e.g., FACT-General total score); and 5) analysis of outcome score (e.g., ANCOVA on post-intervention means adjusting for baseline values) (Figure 1).

**Figure 1.**
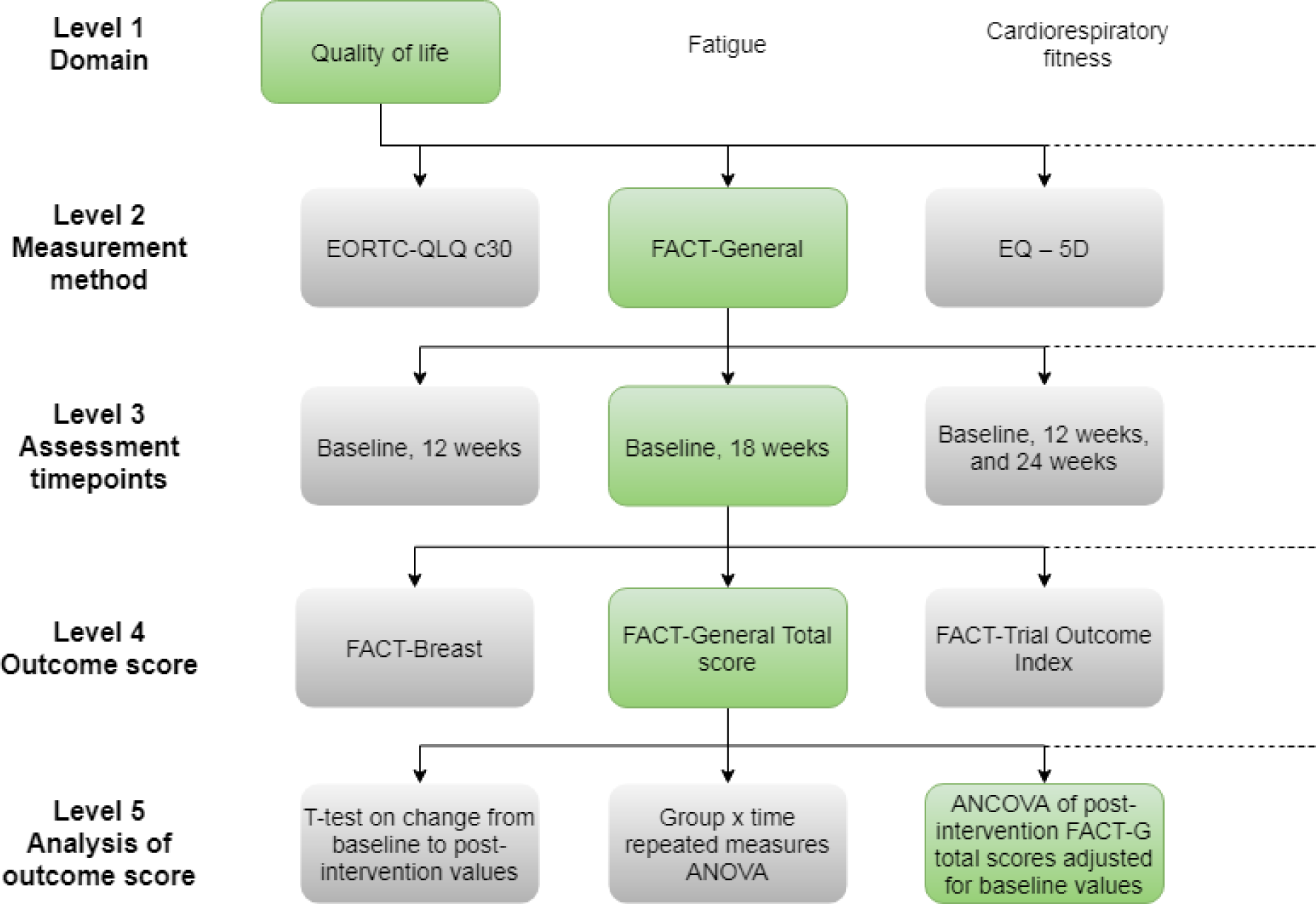
Example of the 5 Levels of Specification in Reporting Outcome Measures. The boxes in green provide an example of sufficient reporting at each level.

If trial authors prespecified only the outcome domain (level 1) but not an outcome score (level 4), we referred to this as an ‘outcome domain-only’ preregistration. Researchers registering their trials on clinical trials registries are directed to provide complete definitions of all outcome measures (scores), under the principle that the information provided should be sufficient to allow other researchers to use the same outcomes. Specifically, Clinicaltrials.gov states that researchers should: 1) state the name of the specific primary or secondary outcome measure (e.g., a descriptive name of the scale, physiological parameter, or questionnaire); 2) provide a description of the metric for how the collected measurement data will be aggregated (e.g., mean change from baseline); and 3) report the timepoint(s) at which the measurement is assessed for the specific metric used. The two examples given for appropriate outcome measure descriptions are: “Number of Participants with Treatment-related Adverse Events as Assessed by CTCAE v4.0,” and “Mean Change from Baseline in Pain Scores on the Visual Analog Scale at 6 Weeks” (46, 47). Similarly, the WHO requires researchers to provide the “name of the outcome, the metric or method of measurement used (be as specific as possible), and the timepoint(s) of interest” (16).

Although a pre-planned statistical analysis is considered an essential component of preregistration (22,23,48), clinical trial registries or the ICMJE (40) do not explicitly require trialists to register their statistical analysis. Therefore, we assessed the completeness of preregistration that would meet 1) WHO/ICMJE standards (i.e. outcome method, score, and assessment timepoints) and 2) the standards that would allow readers to distinguish between analyses planned *a priori* and those planned *post hoc* (i.e. outcome method, score, assessment timepoints, and statistical analysis).

### Analysis of outcomes reported

The decision tree in Figure 2 illustrates how we handled each outcome identified in a prospective registration. First, we distinguished between outcomes prespecified with an outcome score (we defined these as “sufficiently preregistered outcomes”) and those domain-only registered. For preregistered domain-only outcomes, we checked the paper for related-outcomes (e.g., FACIT-Fatigue total score reported as an outcome in a publication would be related to a domain-only entry of ‘Fatigue’ in the linked registry). This represents a necessary deviation from our planned approach for the current study. Our intended and more conservative approach of including only outcomes with prespecified measurement methods, outcome scores, assessment timepoints, and descriptions of how the metrics were calculated and used, would have yielded too few outcomes for our analysis. In addition, this deviation allowed us to account for all outcomes reported in eligible trial publications. For full transparency, we have provided decisions made regarding outcomes in Supplementary Table 2.

**Figure 2.**
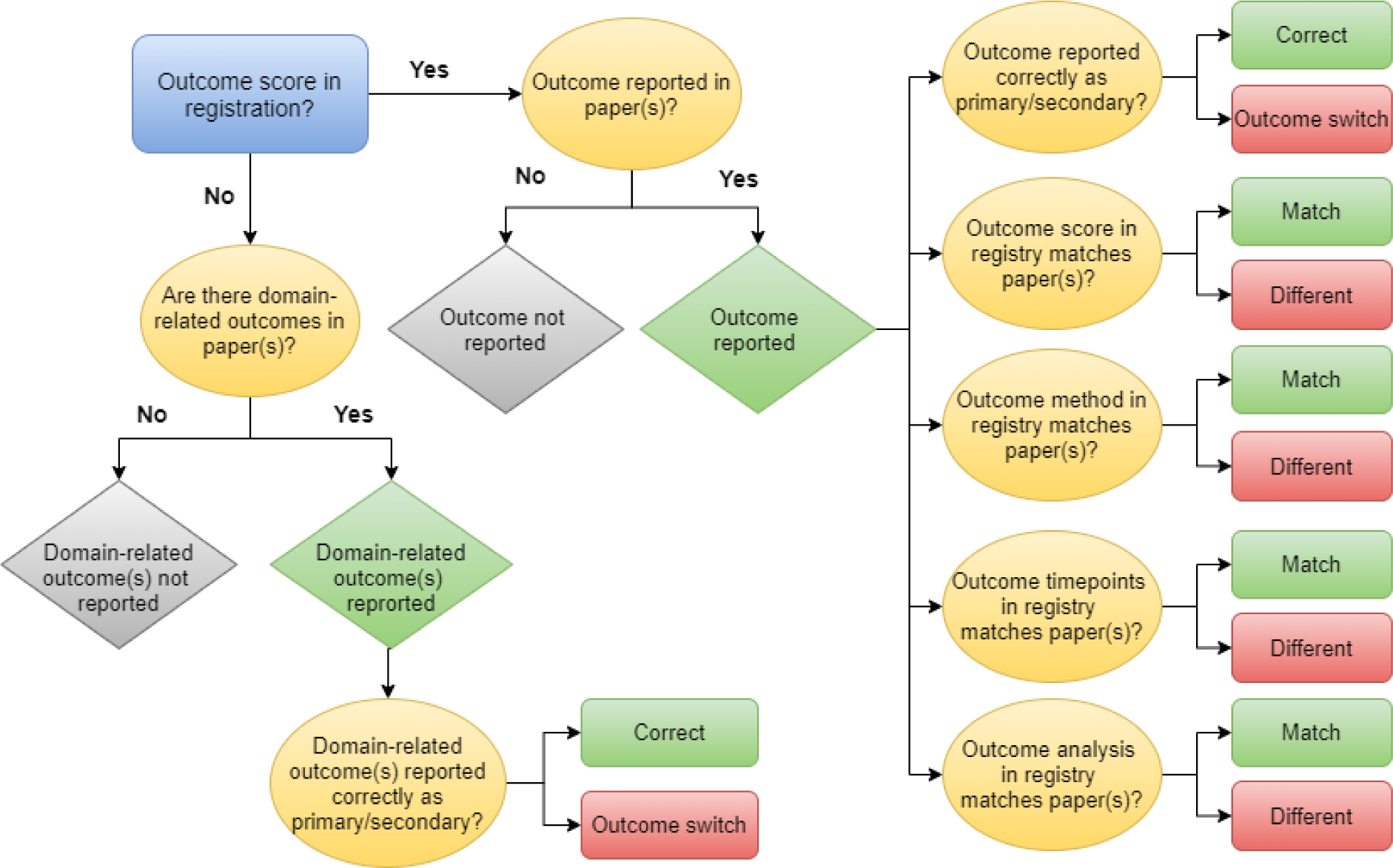
Outcome reporting categorisation decision tree.

For outcomes prespecified with outcome scores in the registry entries, we first recorded whether they were reported in or omitted from trial publications (Figure 2). In the case of the omitted outcomes, we also noted whether they were prespecified as primary or secondary outcomes. Sufficiently preregistered outcomes reported in eligible publications, were then checked to determine whether outcome switching had occurred. In line with the CONSORT guidelines (49), if researchers made any changes to trial outcomes after the trial commenced but clearly reported these changes in the published manuscript, we considered these outcomes to be correctly reported (because the reason for outcome switching was declared and likely to be justified). We reported these cases where present.

For those primary outcomes preregistered with scores, we assessed whether discrepancies—outcome switching and novel outcomes silently introduced as primary outcomes—favoured ‘statistically significant’ results. A discrepancy was considered to favour statistically significant results when: 1) a non-statistically significant (*p*-value > 0.05, or above an *a priori* threshold stated by the authors, or a confidence interval that crossed zero) preregistered primary outcome was demoted to a secondary or non-primary outcome in the published manuscript; 2) a statistically significant (*p*-value <0.05 or below an *a priori* threshold stated by the authors, or a confidence interval that did not cross zero) preregistered secondary outcome promoted to a primary outcome in the published manuscript; and 3) newly introduced non-preregistered statistically significant primary outcomes in the published article. We selected the results of between group comparisons as *a priori* but in cases of within-group comparisons we noted results favouring the exercise group(s) versus control. If we found analyses of the same outcome at more than one timepoint, we noted any comparison favouring the exercise intervention(s) over controls.

We labelled all outcomes reported in published manuscripts not linked to a preregistered outcome score but related to (insufficiently) prespecified domain-only outcomes as ‘domain-related’ outcomes. For all domain-related outcomes, we checked whether the descriptions of outcomes in the published manuscripts matched that of the registries (i.e., primary and secondary domains in registry reported as primary and secondary outcomes in publications, respectively) or whether switching had occurred. If no domain-related outcomes were reported in the available manuscripts, we noted this as ‘domain-only outcomes not reported’.

All outcomes reported in published manuscripts not linked to outcomes preregistered with or without an outcome score, were described as non-preregistered or ‘novel’ outcomes. We noted if authors had declared these ‘novel’ outcomes as non-preregistered (e.g., labelled as an exploratory or secondary analysis), and whether they described them as primary or secondary outcomes in the publications. We considered all undeclared novel outcomes as discrepancies.

### Reporting consistency of preregistered outcomes with scores: clinical registries vs. publications

For outcomes preregistered with scores, we assessed the consistency of reporting in published manuscripts (Figure 2). We noted if the outcome score, outcome method, assessment timepoints, and outcome analysis plan in the preregistration (and published protocols, if available) matched the descriptions in the manuscripts. We considered reporting consistency both with and without the statistical analysis item, due to the absence of this requirement in registry guidance. In additional analyses, we assessed the completeness of statistical reporting for each outcome, and details of this methods and findings can be found in Supplementary Table 3.

## RESULTS

### Study selection

Our PubMed search using Secondary Source IDs resulted in 31 prospectively registered RCTs investigating the effects of at least partially supervised exercise interventions in cancer populations across the selected clinical trial registries (Figure 3). The full list of eligible trials with a reference list for published protocols and results manuscripts are provided in Supplementary Table 4, and a list of excluded trials with reasons is presented in Supplementary Table 5. Most of the 31 trials were registered on Clinicaltrials.gov (n=22; 71%) and the remaining trials were registered on ANZCTR (n=6; 19%), NTR (n=3; 10%; 1 of these 3 trials was also registered on the ISRCTN registry).

**Figure 3.**
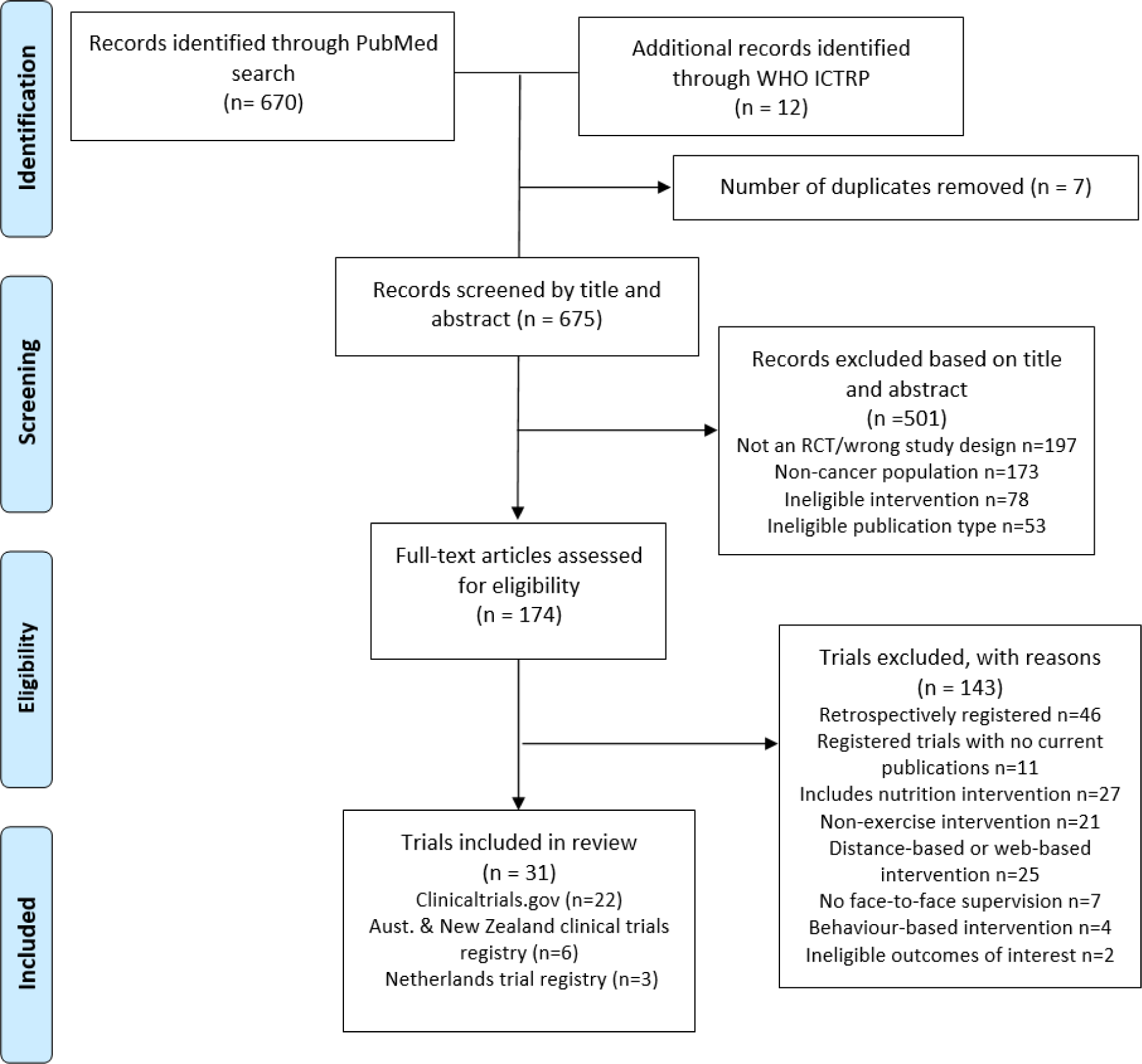
PRISMA diagram of flow of studies.

We found 46 trials that met all eligibility criteria except prospective registration (Supplementary Table 6 and Supplementary Figure 1). These retrospective trials were registered months after trial start dates in most case; there was a median delay of 202 days (IQR =561; min-max = 19-1,770 days;). Most (n=40; 87%) were completed and had published results, however, two were completed but had not published their results yet (last checked: 22^nd^ January 2021), two had been terminated [a reason was given for one: low recruitment (50)], and one had been withdrawn. Only one of the retrospectively registered trials, the large multi-national INTERVAL GAP-4 trial, was currently recruiting (51).^1^

Our search also uncovered 11 trials that were eligible but had not published their results at the time of writing (22^nd^ January 2021) (Supplementary Table 7 and Supplementary Figure 2). Of these 11, we found four completed trials (i.e., actual trial completion dates were provided in their registration). The median number of days from actual study completion to the present day (22^nd^ January 2021) was 366 days, but this time ranged between 205 and 1,075 days. One other trial (52) was suspended (no reason provided). Six of the trials without published results were ongoing trials, and only one had exceeded their anticipated completion date [721 days over as of 22^nd^ January 2020 (53)]. The remaining five studies had a median of 222 (min-max: 38-648) days from the time of writing to the anticipated study completion date (as recorded in the trial registrations).

Trial start dates were often difficult to establish from the registrations. We discovered eight trials on ClinicalTrials.gov that modified their start dates during the study period. Because we did not foresee this issue when writing our preregistration, we devised a *post hoc* solution to establish eligibility. Where recruitment commenced after the start date modification, we accepted the new start date as a prospective registration. However, if a study changed their start date after recruitment had already begun, according to their recruitment status, we rejected the new start date and retained the original start date.

Two trials are noteworthy with respect to the above rule (54, 55). First, Dieli-Conwright (54) initially recruited from May 2010 until the trial was suspended over a year later (14^th^ October 2011). Then on 29^th^ May 2012, the authors made substantial changes to the trial design (new exercise prescription, intervention duration, patient eligibility, and primary and secondary outcomes), before restarting the trial in May 2012. Due to the suspension of the trial in the interim, the author team agreed by consensus to include it in OREO; we therefore, discarded the original registration and accepted the new start date May 2012 was accepted as a prospective registration. However, in another trial (55), recruitment was open for more than two years (August 2012 to 11^th^ January 2015) before the researchers moved the start date forward from August 2012 to December 2014. Because the trial was not suspended and extensive modifications were made to trial design (eligibility criteria, intervention characteristics, and outcomes) in the period between the two start dates, there was a consensus that the new start date should be rejected. However, because the first trial registration was submitted (September 2012) after the original start date (August 2012), the trial was considered retrospectively registered and excluded from OREO. No declarations regarding the modifications to trial start dates, recruitment status, design, or outcomes were made in the published manuscripts for this trial.

### Study characteristics

The characteristics of the 31 eligible trials are presented in Supplementary Table 8. Just over half of the trials (n=18) had published protocols, but only three trials published prospective protocols [NCT03087461 (44, 56); ACTRN12610000609055 (57, 58); ACTRN12611001158954 (43, 59)]. Excluding published protocols, we found a total of 78 published articles across all eligible trials, with a median of two results papers per trial (min-max: 1-7 papers; Supplementary Table 4).

Most trials had small sample sizes, with 17 (55%) trials comprising samples below 100 participants [median (min-max) participants per study: 65 (23–420)]. The most commonly studied cancer type across trials was breast (n=13; 42%) followed by prostate (n=6; 19%).

Participants were aged on average 58 ± 8 years (mean ± SD) across trials and had an average BMI in the ‘overweight’ category (mean ± SD: 28 ± 3 kg/m^2^). Based on thirteen (42%) trials with available ethnicity data, participants were mainly white (mean ± SD: 72 ± 26%). From the 26 (84%) trials with cancer stage data, most patients had a diagnosis of either stage II or III cancer (57%). Most trials (n=17; 55%) comprised patients who had completed adjuvant therapy (chemotherapy or radiotherapy), whereas 12 (39%) trials consisted of patients receiving either chemotherapy or radiotherapy (n=9), androgen deprivation therapy (n=2), or aromatase inhibitors (n=1), and two (6%) studies included patients awaiting surgery.

Most exercise interventions comprised both aerobic and resistance training (n=14; 45%), or aerobic exercise alone (n=9, 29%). Interventions were relatively short-term on average (mean ± SD: 18 ± 13 weeks), and the two longest trials both lasted a year (60, 61). We found five (16%) partially supervised trials (less than half of prescribed exercise sessions were supervised in-person), whereas all others were fully supervised (n=26; 84%).

### Completeness of preregistration

All decisions made regarding the included trials can be accessed here: https://bit.ly/3qFkcro; and our raw data can be found here: https://osf.io/4c8mb/. In the 31 eligible trial registrations, we found a total of 405 prespecified outcomes: 71 were recorded as primary, and 334 were secondary outcomes (Figure 4). There were a median of 11 outcomes per registration (min-max: 1-59), and the median time between registration and trial start was 28 (0–785) days (Supplementary Figure 3).

**Figure 4.**
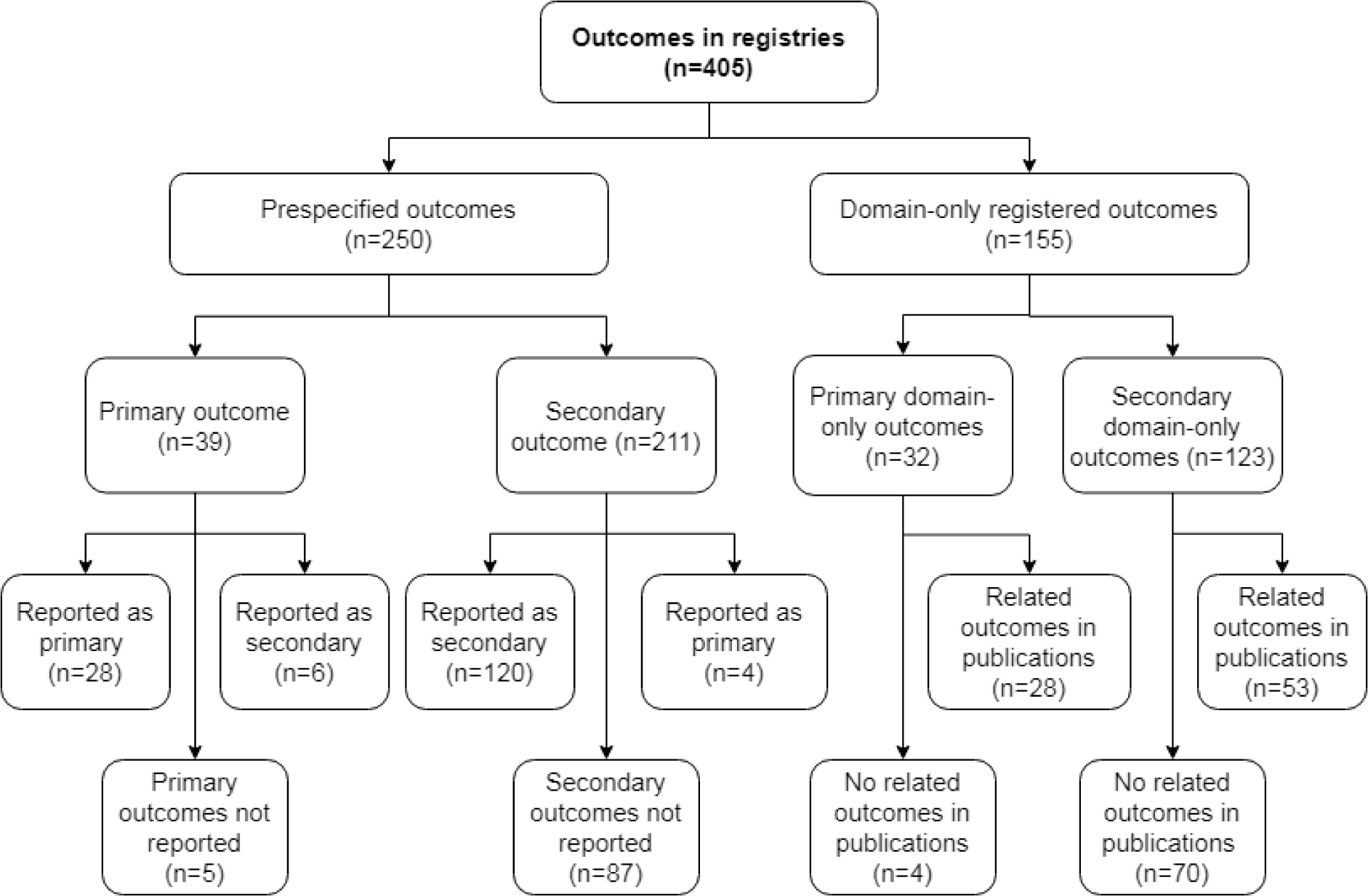
Summary of the fates of outcomes reported in trial registries.

The completeness of preregistration varied across trials (Table 2 and Supplementary Figures 4-7). Only 25 of a total 405 outcomes (6.2%) were preregistered completely, that is, the registration provided an outcome score with a method of measurement, assessments timepoints, and a planned statistical analysis description. These 25 outcomes came from just three studies (56,58,62). If we disregard a planned statistical analysis description as a necessary element of preregistration, then still less than half of the outcomes would be completely preregistered (n=175; 42.5%) and more than one-third of trials would have provided no preregistered outcomes (n=12; 38.7%; Table 2).

**Table 2.**
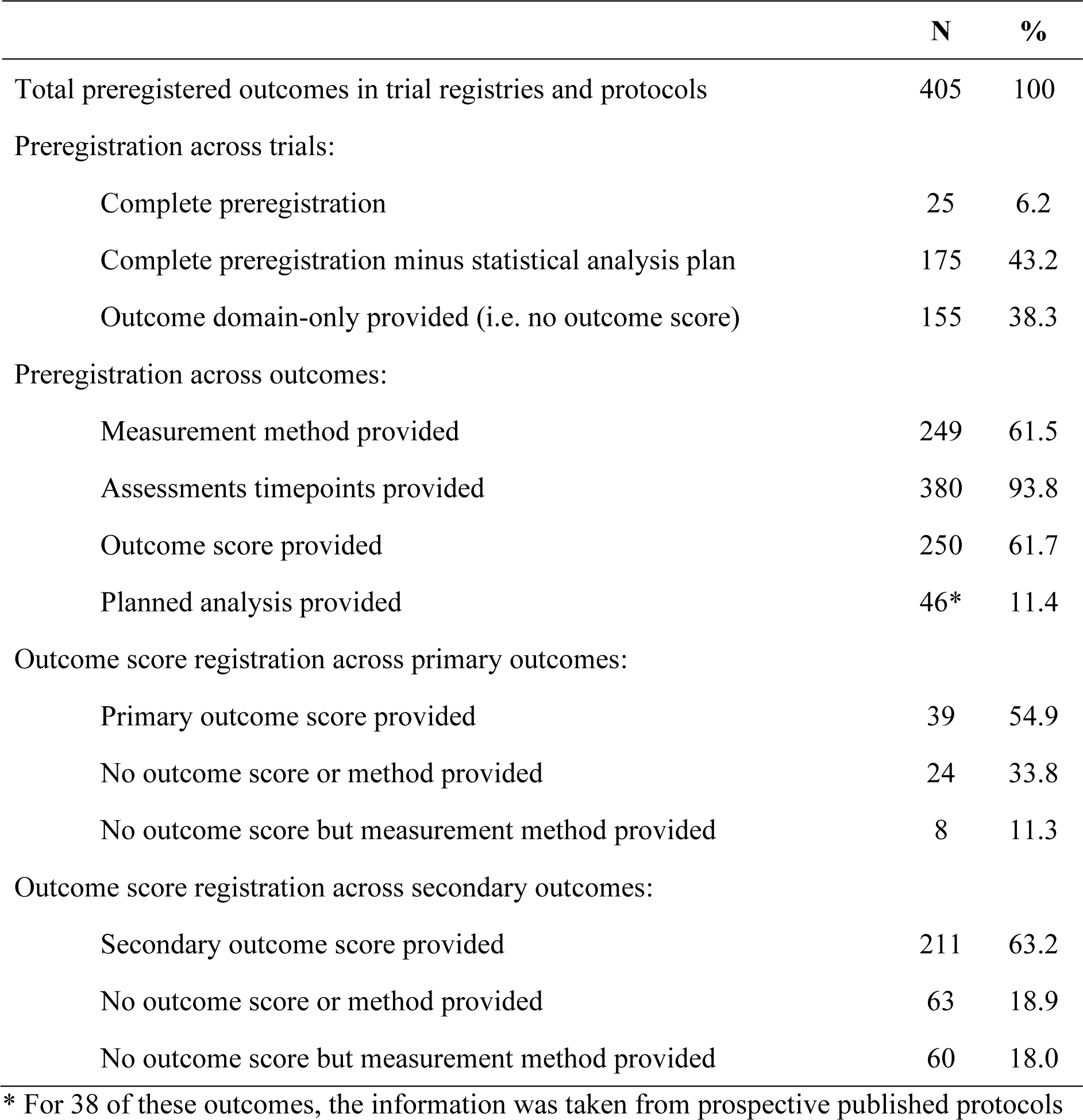
The completeness of outcome preregistration.

Assessment timepoints were provided for most preregistered outcomes (n=380, 93.8%), with 25 (80.7%) trials specifying timepoints for all outcomes listed, but four (12.5%) trials failed to report assessment timepoints for any listed outcome. However, only 61.5% of the 405 (n=249) preregistered outcomes specified a method of measurement. Only nine (29.0%) trials preregistered a measurement method for all their outcomes, whereas another 10 trials did not provide any measurement method in their registration.

Researchers preregistered an outcome score for 250 (61.7%) outcomes (Table 2). Six studies (19.3%) provided no outcome scores for any of their outcomes, and therefore, included no sufficiently preregistered outcomes (63–68). Only two (6.5%) trials provided an outcome score for all preregistered outcomes. Of the 155 (38.3%) outcomes where no score was preregistered, 68 (16.8%) provided a measurement method and 87 (21.5%) provided only the outcome domain with no method. A slightly higher percentage of preregistered primary outcomes were missing an outcome score than secondary outcomes (33.8% vs. 18.9%).

A planned statistical analysis was preregistered (either in registry entries or prospective published protocols) for 46 (11.4%) outcomes across three studies (30 from one trial: (58); 8 each from 2 studies: (56, 62)]. These three studies provided an analysis plan for all their outcomes. We found the statistical analysis plan for nine (2%) of outcomes in the registry entries (from two trials) and obtained the remaining preregistered analysis plans from prospective published protocols. One other trial reported two possible statistical approaches (two-way, group x time repeated measures ANOVA or ANCOVA “as appropriate”, with no elaboration), and we, therefore, marked this study as not having stipulated a specific planned statistical analysis (59).

### Reporting of outcomes prespecified with an outcome score in publications

Outcomes prespecified in a clinical registry with an outcome score and reported in publications without outcome switching represented just 15.9% (148/929) of all published outcomes (i.e. outcomes not omitted, prespecified primary outcomes reported as primary and prespecified secondary outcomes reported as secondary in publications).

When we considered only outcomes preregistered with an outcome score, we found that most (59.2%; n=148/250) were published without switching or omission (Figure 4). More than a fifth (22.7%; n=92) of all preregistered outcomes with an outcome score, however, were not reported in publications, and 2.5% (n=10) were switched (9 were switched without declaration). Almost a third (n=10/32) of trials failed to report a single sufficiently preregistered outcome that was not switched or omitted from their published manuscripts.

We identified 39 sufficiently prespecified primary outcomes (i.e. with an outcome score). Of these, the authors reported 28 (71.8%) correctly as primary outcomes in the published papers (Figure 3), omitted five (12.8%) from published manuscripts [one study (69) provided their finding in the results section of the registry; the finding was not statistically significant], and reported six (15.4%) sufficiently prespecified primary outcomes as secondary outcomes in the published papers (i.e. outcome switching). However, for one of the primary outcome switches, the authors declared the switch and provided that the following reason: “the need to reduce the study’s sample owing to a reduction in funding” (60). Therefore, we found five cases of undeclared outcome switches in primary outcomes with prespecified outcome scores. Taken together, 25.6% (n=10/39) of sufficiently prespecified primary outcomes were switched for a secondary outcome without declaration or omitted from published manuscripts (secondary aim 1).

Researchers reported 120 (56.9%) of the 211 secondary outcomes that provided an outcome score correctly as secondary without switching or omission (secondary aim 2). We discovered a relatively large number of unreported sufficiently preregistered secondary outcomes (40.7%; n=87), and four (1.9%) cases of undeclared outcome switching (i.e. prespecified secondary outcomes were reported as primary outcomes; Figure 4).

### Reporting of domain-only outcomes

We could link 81 (52.3%) outcomes preregistered as ‘domain-only’ (e.g., only ‘fatigue’ prespecified as an outcome in the registry and FACT-Fatigue total score reported in a linked publication) to published outcomes (Figure 4). Most (87.5%; n=28/32) primary outcomes that were preregistered without an outcome score (domain-only) had related outcomes in the published papers, whereas, a minority (43.1%; n=53/123) of secondary-domain-only outcomes related to published outcomes.

The remaining 47.7% (n=74) of the outcomes preregistered with only a domain were not related to any published outcome (i.e. omitted outcomes). Most (56.9%; n=70/123) domain-only outcomes dropped from publications were secondary outcomes; conversely, only 12.5% (n=4/32) of primary domain-only outcomes were omitted (Figure 4). Adding these unreported outcomes to the 92 omitted preregistered outcomes with outcome scores (described above), means that 41.0% (n=166) of all outcomes provided in registrations were omitted from the available published manuscripts (Figure 4 and Table 3). This omission of preregistered outcomes is suggestive of selective outcome non-reporting.

**Table 3.** Discrepancies in outcomes and omitted preregistered outcomes across eligible trials.

| Outcomes | N | % |
| --- | --- | --- |
| Total outcomes in eligible publications | 929 | 100 |
| Outcomes labelled as primary outcomes in publications | 170 | 18.3 |
| Outcomes labelled as secondary outcomes in publications | 759 | 81.7 |
| Discrepancies in outcomes |  |  |
| Total discrepancies in outcomes labelled as primary | 84 | 49.4 |
| Primary outcomes in publications that were preregistered as secondary outcomes with an outcome score (undeclared) | 4 | 2.4 |
| Primary outcomes in publications that were preregistered as secondary outcomes without outcome scores (undeclared) | 20 | 11.8 |
| Undeclared novel outcomes published as a primary outcome | 60 | 35.3 |
| Total discrepancies in outcomes labelled as secondary | 395 | 52.0 |
| Secondary outcomes in publications that were preregistered as primary outcomes with an outcome score (undeclared) | 5 | 0.7 |
| Secondary outcomes in publications that were preregistered as primary outcomes without outcome scores (undeclared) | 56 | 7.4 |
| Undeclared novel outcomes published as a secondary outcome | 334 | 44 |
| Total discrepancies across all outcomes (% published outcomes) | 479 | 51.6 |
| Total undeclared outcome switches | 85 | 9.1 |
| Total undeclared novel outcomes | 394 | 42.4 |
| Preregistered outcomes omitted from publications |  |  |
| Total preregistered outcomes omitted from publications (% of preregistered outcomes) | 166 | 41.0 |
| Omitted preregistered primary outcomes | 9 | 2.2 |
| Omitted preregistered secondary outcomes | 157 | 38.8 |
| Number of trials with discrepancies and omitted preregistered outcomes: |  |  |
| Trials with no discrepancies (i.e. no undeclared switching or novel outcomes) | 4 | 12.9 |
| Trials with no omitted preregistered outcomes | 8 | 25.8 |
| Trials with no discrepancies (i.e. no undeclared switching or novel outcomes) or omitted preregistered outcomes | 1 | 3.2 |

Many of the outcomes preregistered without an outcome score were related to several outcomes in publications, therefore, we found that over a third (35.3%; n=328/929) of outcomes in publications were linked to these domain-only outcomes (secondary aim 3A). Of these outcomes, 22.9% (n=75) were reported as primary outcomes and 77.1% (n=253) as secondary outcomes in the published manuscripts. However, we found evidence of outcome switching among these outcomes. More than a quarter of outcomes (26.7%; n=20) reported as primary outcomes in eligible publications were related to domain-only registered secondary outcomes. Similarly, 22.1% (n=56) of outcomes reported as secondary in published articles were related to primary outcomes preregistered as domain-only (Figure 5 and Table 3). Overall, 8.2% (n=76/929) of outcomes in published manuscripts were instances of outcome switching among domain-only preregistered outcomes (secondary aim 3B). An exploratory analysis revealed that the odds of a domain-related outcome being switched was almost four times greater than that of an outcome preregistered with an outcome score (Odds Ratio 3.97; 95% CI 2.12 to 8.16).

**Figure 5.**
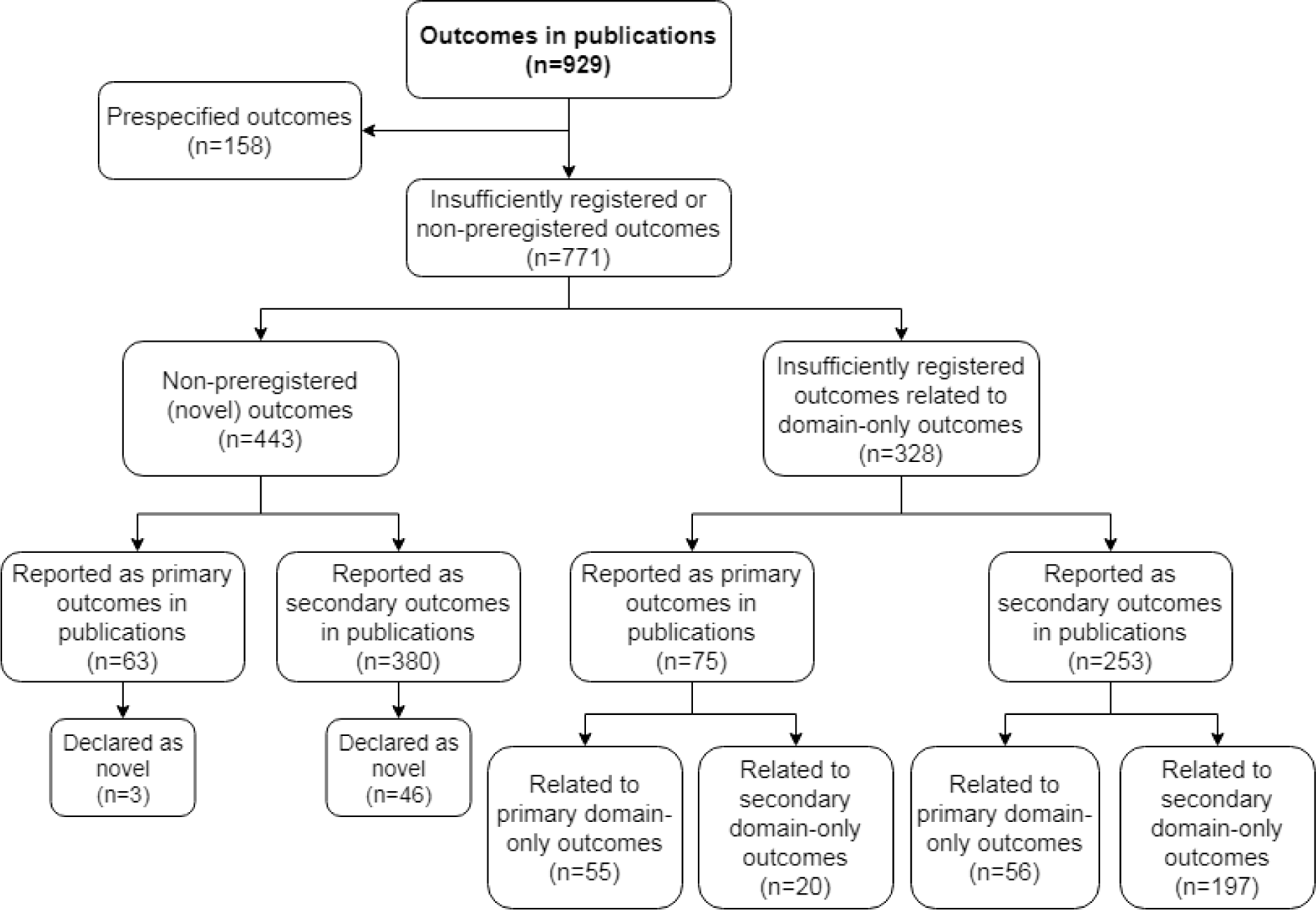
Summary of outcomes published in eligible publications.

**Figure 6.**
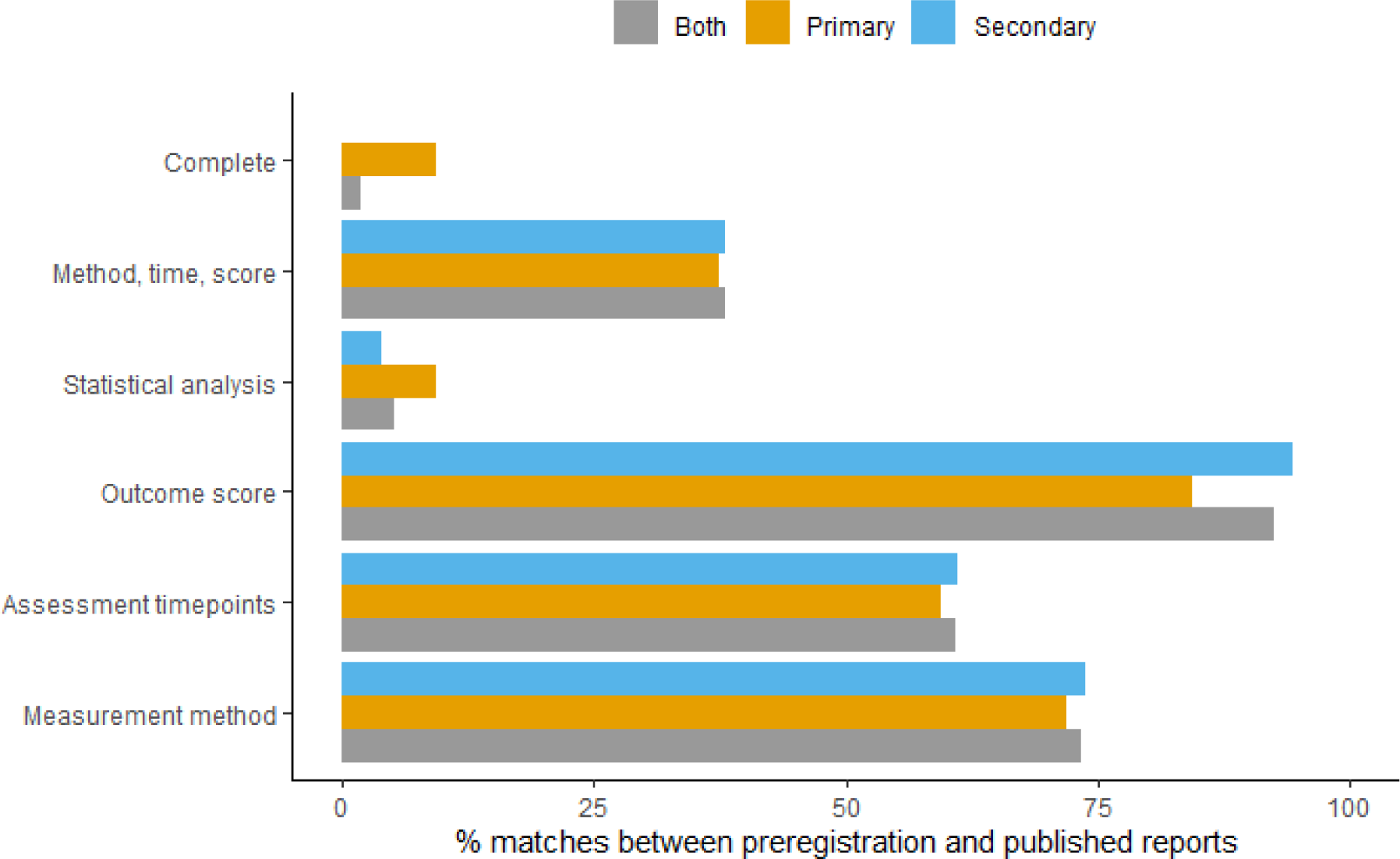
Percentage of sufficiently registered outcomes where the preregistered descriptions of measurement method, assessment timepoints, outcome score, and statistical analysis match the description in the published manuscripts. ‘Method, time, score’ = descriptions of preregistered measurement method, assessment timepoints, and outcome score matches published manuscript. ‘Complete’ = all preregistered elements of the outcome description match the published manuscript. ‘Both’ = both primary and secondary outcomes.

### Non-preregistered (novel) outcomes reported in eligible published manuscripts

Of the 929 published outcomes, only 17.1% (n=158) comprised outcomes prespecified with an outcome score and 35.3% (n=328) were outcomes linked to domain-only outcomes in preregistrations, therefore, most published outcomes (47.7%; n=443/929) were novel outcomes (i.e. not related to any prespecified outcome) introduced after trial registration (Figure 5 and Table 3). Authors reported 14.2% (n=63) of these novel outcomes as primary outcomes, and 85.8% (n=380) as secondary outcomes. Only 11.1% (n=49/443) of novel outcomes were declared as such (mostly by labeling the analysis as ‘secondary’, ‘exploratory’, or ‘post hoc’) (secondary aim 4). Therefore, silently introduced (undeclared) outcomes represented 42.4% (n=394; 61 primary and 335 secondary) of published outcomes (secondary aim 5; Table 3). A median (min-max) of 10 (0–50) novel outcomes were silently introduced per trial.

### Summary of discrepancies in outcomes published in eligible trial manuscripts

Overall, almost one in every 10 (9.1%) outcomes in published eligible trial manuscripts were switched outcomes without declaration, and 42.4% were undeclared novel outcomes (Table 3 and Supplementary Table 9). Half (49.4%) of the outcomes labelled as primary outcomes in published manuscripts were switched or undeclared non-preregistered outcomes (Table 3). Therefore, taken together, half (50.7%) of all published outcomes were either switched or silently introduced post hoc. Twenty-seven (87.1%) of the 31 trials contained at least one discrepancy. Eight trials (25.8%) omitted preregistered outcomes. Only one (3.2%) trial was free from switched, undeclared novel, or omitted outcomes [registration: (70); publication: (71)].

Of the 152 correctly reported outcomes, 38 (25%) were described as statistically significant. All four secondary outcomes promoted to primary outcomes in published manuscripts were statistically significant. However, all five primary outcomes demoted to secondary outcomes in published manuscripts also attained statistical significance. Of the 50 undeclared novel efficacy-related outcomes introduced as primary outcomes in publications, 22% (n=11) were statistically significant.

### Proportion of published prespecified outcomes descriptions matching trial results publications

We examined the consistency of reporting between preregistration and published outcomes by comparing the reporting of the outcome measurement method, assessment timepoints, outcome score, and statistical analysis of sufficiently prespecified outcomes (i.e. with an outcome score) in prospective registrations to the published manuscripts. Of the 158 (primary: 34; secondary: 124) prespecified outcomes in published manuscripts, the described preregistered measurement method matched the published report for 116 (73.4%) cases, whereas the assessment timepoints and outcome score were consistent for 96 (60.8%) and 146 (92.4%) cases, respectively.

There were 62 (39.2%) mismatches between assessment timepoints reported in registry entries and published manuscripts, and most of these mismatches were due to omitted timepoints (74.2%; n=46/62). For the remaining inconsistencies, researchers introduced extra timepoints for six (9.7%) outcomes (e.g., a non-preregistered follow-up assessment was added), reduced the time between assessments for three (4.8%) outcomes, and extended the timepoints for one (1.6%) outcome (e.g., a preregistered post-intervention assessment for 12 weeks reduced to eight weeks or extended to 18 weeks in publication). For six (9.7%) outcomes the preregistered assessment timepoints were unclear. When a pre-planned statistical analysis was available, only eight (5.1%) of them matched that reported in the published protocol papers.

For the 25 completely preregistered outcomes (described above and in Table 3), the published description of only three outcomes matched the trial registry entry (secondary aim 6). These three outcomes came from one study (56) and were descriptive analyses of feasibility outcomes (recruitment, retention, and adherence). Therefore, we found no preregistered efficacy or effectiveness outcomes that were measured and assessed as planned when we consider prespecification of measurement method, outcome score, assessment timepoints, and planned statistical analysis as essential components of preregistration.

If we exclude statistical analysis plans, descriptions of measurement method, assessment timepoints, and outcome scores in the clinical trial registrations matched that reported in published manuscripts for only 60 (38% of 158 sufficiently preregistered outcomes; and 6.5% of published outcomes) outcomes (only 12 of these were primary outcomes).

All descriptions of preregistered outcomes with a score matched that in the published report in just four trials (72–75); however, these trials also comprised insufficiently preregistered (domain-only) outcomes, omitted outcomes, and undeclared novel outcomes. Therefore, no trial was free of inconsistencies (i.e. insufficiently described prespecified outcomes, unmatched outcome descriptions, switched outcomes, and undeclared novel outcomes).

## DISCUSSION

Our current analysis aimed to evaluate selective outcome reporting in exercise oncology RCTs by comparing prospective trial registrations with their associated published articles. We found trial registrations lacking essential details, and widespread evidence of outcome switching and reporting bias. None of the 78 published articles from the 31 trials were free from incomplete, inconsistent, or selectively reported outcomes. Fewer than 20% of outcomes reported in published manuscripts were sufficiently preregistered and presented without switching, and 42% of published outcomes were silently introduced (undeclared) novel outcomes. Furthermore, when we compared descriptions of sufficiently prespecified outcomes in registry entries to published manuscripts, we found inconsistencies for all but three outcomes from one study. These findings raise questions about the integrity of the exercise oncology literature.

### Prospective and retrospective trial registrations

Despite the ICMJE (18), WHO (19), and the Declaration of Helsinki (17) mandating that every clinical trial is registered in a publicly accessible database before recruitment of the first participant, we discovered 46 otherwise eligible exercise oncology RCTs that were retrospectively registered. Although these retrospectively registered trials in our search results started years after the launch of such requirements (100% were registered in or after 2005; 80% were registered in or after 2008), most were registered months or even years after the first patient was enrolled. Therefore, more than 50% of trials that met all other eligibility criteria were not compliant with international standards [e.g. WHO and WMA (19)] for clinical research.

### Outcome switching, omission, and silent introduction

The most critical type of discrepancy involves the switching of a preregistered primary outcome for a non-primary outcome in the published article. Researchers select the primary outcome in advance as the most clinically relevant measure that addresses the trial’s main aim. As a rule, sample size justifications are based on the sample size required to detect an effect on the primary outcome (76). Changing the primary outcome threatens the validity of a study and can lead to an overestimation of intervention effects (77). Therefore, it is worrying that half (n=84) of the 170 outcomes labelled as ‘primary’ in published manuscripts were either undeclared promotions of outcomes preregistered as secondary outcomes or silently introduced novel outcomes. Only 28 outcomes labelled as ‘primary’ in published manuscripts were prespecified with an outcome score in a registry entry or protocol paper, and another 55 could be linked to a domain-only preregistered outcome.

We observed similar practices in secondary outcomes: 395 (52.0%) of the 759 outcomes labelled as ‘secondary’ in published manuscripts were undeclared switches or silently introduced novel outcomes. Just 15.8% of outcomes labelled as ‘secondary’ in publications were preregistered secondary outcomes with scores, whereas another 26.0% were related to preregistered secondary domain-only outcomes. Switching and omission were widespread and were absent from only four of the eligible trials.

Of note, 41% of all preregistered outcomes were omitted from published manuscripts. Researchers, however, omitted secondary outcomes more often than primary outcomes (38.8% vs. 2.2%). Only eight trials had no omitted outcomes, and just one was free of any discrepancies (i.e. no undeclared switching or novel outcomes) or omitted preregistered outcomes. Similarly, most trials (n=26) silently introduced novel outcomes in the published results manuscripts. These novel outcomes accounted for 42% of all outcomes reported in the manuscripts, with a median of 14 outcomes silently introduced per trial.

Our findings are consistent with other studies that have explored discrepancies between registry entries and published manuscripts. A 2008 systematic review of studies exploring outcome reporting bias in RCTs of healthcare interventions discovered that one or more primary outcomes were changed, introduced, or omitted in 40–62% of studies (9). A later systematic review in 2015 detected a similar proportion of trials [median 31%; IQR 17–45%] with a discrepancy between the registered and published primary outcome (24). Goldacre and colleagues (26) observed that outcome misreporting is also common in top medical journals that endorse the CONSORT statement, with outcome discrepancies requiring a correction letter identified in 87% of included trials.

Deviations from preregistered protocols are a normal part of the scientific process and are not unexpected when the process involves a complex behavioural intervention, such as exercise, and a heterogeneous population like patients with cancer. Considering the volume of discrepancies between preregistered and published outcomes we observed, surprisingly, only one study transparently declared an outcome switch (78). When researchers do not provide such acknowledgements, one cannot rule out the possibility that the researchers’ preference towards a certain (often favourable) finding motivated the decision to omit outcomes and not declare discrepancies between the preregistration and the published article. One major incentive for such outcome switching and selective outcome reporting is publication bias—the historical preferential publishing of positive findings and studies that find support for their hypothesis (79). Regardless of the motivation, however, failure to declare such deviations contravenes the Declaration of Helsinki and leads to published articles that are dishonest, misleading, and potentially harmful to patients (80).

### Vague preregistration and inconsistent reporting of outcomes

Authors of clinical trial registrations must describe their primary and secondary outcomes in sufficient detail (akin to reporting enough detail in a methods section to allow replication by independent researchers). For example, describing an outcome simply as ‘fatigue’ (domain only) is vague and would be unacceptable in a scientific article. In exercise science, fatigue can refer to both a subjective experience and an objective change in task performance. In the present context, fatigue refers most often to a construct that is measured as a patient-reported outcome, but many validated questionnaires exist for this purpose, and some include both sub-scales and a total score. Therefore, when vague terms such as ‘fatigue’ are used, readers have no way of ascertaining which questionnaire and score the researchers originally selected to assess this outcome. Unfortunately, we found many such examples of poor reporting quality across the 31 trial registrations, which hampered our task of evaluating reporting bias. We found that 35.3% of outcomes were provided without outcome scores (domain-only), and six studies (19.4%) provided no scores for any of their outcomes, and therefore, included no sufficiently preregistered outcomes.

We evaluated outcome preregistration on five levels: the domain, measurement method, outcome score, assessment timepoint, and planned statistical analysis [modified from Zarin et al. (45)]. We found just 26 (6% of outcomes in trial registrations) outcomes that included all five levels (i.e. completely preregistered outcomes), and authors accurately reported only three of these outcomes [all from one trial (56)] in the published article (i.e. descriptions in publications matched the registration). Unfortunately, none of the outcomes were health-related outcomes— all three outcomes were feasibility-related.

When we excluded the requirement of a planned statistical analysis (which is usually a requirement for institutional review board review and approval, but not an element for clinical trial registries), we found that less than half of the preregistered outcomes were reported with all other components, and only 38% were reported consistently in the published article (i.e. a match between registration and publication outcome descriptions). Only 12 of these outcomes were primary outcomes, and of these: one was switched to a secondary outcome in the published report [in trial (81)], three were feasibility outcomes [mentioned above from the Bridging the Gap trial (56)], and only eight were efficacy outcomes reported correctly, albeit without a prespecified analysis plan. Interestingly, the statistical analysis of only three of the eight primary outcomes produced a *p*-value below 0.05 (the most common threshold for ‘statistical significance’) in favour of the intervention group. We identified no trials free from outcome switching, omission, or misreporting.

Although researchers could provide an ambiguous description of outcomes unintentionally, the result is that it masks the researcher’s degrees of freedom at the point of analysis and dissemination. Vague outcome descriptions bestow researchers with the flexibility to choose their preferred measurement of the construct (e.g. if two fatigue questionnaires have been included), metric or scoring method (e.g. prioritizing a specific subscale) and assessment timepoint. Moreover, when researchers do not provide a planned statistical analysis in advance, they have greater flexibility to analyze the data in many ways using different statistical approaches and models. Researchers can then select, often based on the direction, size or statistical significance (e.g., *p*<0.05) of an effect, the approach or model that produced their favoured result and discard less favourable ones. Although this undisclosed flexibility in outcome and analysis selection may increase the likelihood of publishing, it inflates the Type 1 error rate to an unknown extent and increases the likelihood that the preferred ‘positive’ findings presented in the published article occurred by chance alone. In other words, published articles may are more likely to include false positives and exaggerate the benefits of an exercise intervention.

Exercise is considered medicine in oncology (82), but for key stakeholders to appropriately recognize exercise as adjunct care, exercise RCTs must be conducted and reported as intended—to minimize bias. RCT design seeks to provide high-quality evidence about the causal relationship between an intervention and an outcome by lowering harmful biases to a greater extent than observational studies (83). Important RCT elements such as random sequence generation and allocation concealment (selection bias) and the blinding (masking) of trial personnel and participants (performance and detection bias), when applied appropriately, can increase the likelihood the study’s findings are accurate estimates of the true effect (84). However, flaws in the design, conduct, analysis, and reporting of RCTs can undermine their ability to yield reliable causal inferences and produce underestimates or overestimates of the true intervention effect (i.e. bias) (85). Efforts to minimize biases in trial design and conduct are futile if the subjective influence of the researcher is embraced at the point of analysis and dissemination (reporting bias). Reporting bias in exercise oncology trials undermines not only individual RCTs, but also any systematic reviews with meta-analyses for which the individual RCTs are eligible. Furthermore, the credibility and certainty of position stands and clinical recommendations (4,7,86,87) may be weakened if they are informed by evidence afflicted with reporting bias.

Although prospective trial registration did not appear to prevent reporting bias, it has allowed us to evaluate the existence of this bias in the exercise oncology literature. We encourage those wishing to evaluate the robustness of a finding reported in a registered exercise oncology trial publication by 1) checking that the outcome of interest is present, described in sufficient detail, and prospectively registered by inspecting the history of revisions record in the clinical trial registration, and 2) scrutinising the published manuscript for any declaration of a discrepancy (e.g. a reason why the outcome is missing or switched) compared with the trial registration. Prospective registration of a clinical trial registry is not yet a sufficient deterrent against reporting bias, therefore, alongside raising awareness about this issue, other solutions should be considered.

### The Solutions

Clinical trials (see ICMJE definition) involving exercise for people with cancer must be prospectively registered before recruitment of the first participant. Prospective trial registration is not optional for researchers, they have a scientific and ethical responsibility to participants to comply, and we call for funders, editors, article reviewers, researchers and other stakeholders in exercise oncology to consider it mandatory. Specifying outcomes in sufficient detail (all 5 levels of outcome specification in Figure 1) is paramount; we observed a higher prevalence of outcome switching in outcomes described without an outcome score in trial registrations. Transparent reporting and justifying deviations from the registered protocol (in Supplementary Material or an open archive) is a normal part of the scientific process and should be adopted as the standard by exercise oncology researchers (as recommended by CONSORT guidelines; http://www.consort-statement.org/). Without such detail, readers can not easily establish which outcome analyses are confirmatory or exploratory in published articles.

Trial registrations serve important functions for promoting the fulfillment of ethical obligations to participants and the research community, such as providing information to potential participants and referring clinicians and providing a public record of study outcomes and results. However, conceptually preregistration allows independent researchers to transparently evaluate the capacity of a trial or test to falsify a prediction [the severity of a test; see Lakens (88)]. This independent evaluation is not possible if the clinical trial registration is lacking key aspects in comparison to the more comprehensive preregistration standards that have arisen from the replication crisis in psychology (89–91). The absence of statistical analysis plans from most of the trial registrations included in our sample precludes an assessment of how severely the predictions they made were tested. Therefore, currently, most of the exercise oncology trials may only provide what Professor Deborah Mayo, one of the chief proponents of severity testing, describes as ‘bad evidence, no test’ (92). One potential solution to improve preregistration quality is to preregister using SPIRIT reporting guidelines [www.spirit-statement.org; (93)] and platforms that guide and support detailed preregistration such as the Open Science Framework (www.osf.io) and AsPredicted (www.aspredicted.org).

Considering the (often public) investment of resources and participant time and effort, the field of exercise oncology should also consider adopting the registered report format (94). Registered reports are a publishing model that seeks to address selective reporting bias and publication bias (79). The key innovation with registered reports is that peer-review is split into two stages, the first stage takes place *before* participant enrollment, whereas the second occurs after the authors complete the study and submit the final report (for a primer related to exercise science, see Caldwell et al. (95)). The importance of the research question and the validity and rigour of the proposed methods and analysis plan can be assessed (and strengthened) in advance during the first stage. If the reviewers agree that the question is important and the methodology to test it is appropriate, then an “in-principle acceptance” can be granted. This means that the results will be published regardless of the outcome (null or otherwise), assuming the researchers followed their protocol, declared any deviations, and interpreted the results according to the evidence. Registered reports, like preregistration, can also help limit undisclosed analytic flexibility (the garden of forking paths) because they include an analysis plan (91, 94). *BMC Medicine* became the first clinical research journal to offer the registered report format in 2017, and the first registered report in exercise oncology will signify a welcome step for the field toward increased transparency.

### Limitations and Future Directions

We excluded trial registrations that included unsupervised exercise interventions and exercise combined with other intervention components. Therefore, our analysis is not necessarily representative of the exercise oncology literature more broadly, though we have little reason to suggest that the inclusion of these studies would alter our main findings. We could not include 11 trials because they lacked published articles at the time of our search dates. Given that these are more recent trials, we will note with interest if outcomes published in future articles are more closely aligned with their trial registrations.

During our analysis, we made several additional decisions regarding the preregistration of outcome scores were made that were not prespecified in our preregistered protocol; these decisions are outlined with a rationale in Supplementary Table 2. Aside from additional decisions; our primary protocol deviations were as follows: we did not consider each time point as a separate outcome (instead we noted whether assessments for each outcome were reported at each timepoint); we excluded cross-sectional and mediation analysis publications from our main analyses (although this was perhaps explicit from our aim to study ‘intervention effects’); we used multiple authors for data entry (instead of a single author); and we added an exploratory analysis investigating the odds of outcome switching for a domain-related outcome versus an outcome preregistered with a score (Supplementary Table 10). Furthermore, during the data entry process, we introduced a process for the categorisation of the level of outcome registration (as either “outcome domain only”, “partial” or “complete”) to allow us to identify the sources of selective outcome reporting and switching. We planned several secondary analyses that arose from the data that were not preregistered and will be reported in subsequent articles.

## CONCLUSIONS

Half of otherwise eligible exercise RCTs in people diagnosed with cancer were retrospectively registered and, therefore, did not meet current international standards for clinical trials. Across 31 prospective trial registrations, we found evidence suggestive of widespread selective outcome reporting and non-reporting bias. The existence of such outcome reporting bias has implications for the integrity and credibility of randomized trials in exercise oncology. The omission of potentially non-statistically significant, ‘negative’, or small effects in favour of more ‘positive’ or novel findings could be distorting the exercise oncology literature, leading to potentially inaccurate claims about the potential benefits or harms of exercise for people with cancer. Sufficient trial preregistration along with transparent reporting of outcomes and deviations from trial protocols in future exercise oncology trials is warranted to better understand the role of exercise in cancer care.

## AUTHOR CONTRIBUTIONS

Contributor Roles Taxonomy (CRediT: http://credit.niso.org):

| Task | Responsibility of |
| --- | --- |
| Conceptualization | BS (Equal), CMF (Equal), JFC (Equal), KAB (Equal), RT (Support), DN (Equal), IML (Equal) |
| Data Curation | BS (Equal), CMF (Equal), KAB (Support), RT (Support), DN (Support), IML (Equal) |
| Formal Analysis | BS (Support), IML (Lead) |
| Funding Acquisition | Not applicable |
| Investigation | BS (Equal), CMF (Equal), KAB (Support), RT (Support), DN (Support), IML (Equal) |
| Methodology | BS (Equal), CMF (Equal), JFC (Equal), KAB (Equal), RT (Equal), DN (Equal), IML (Equal) |
| Project Administration | BS (Lead), IML (Support) |
| Resources | BS (Support), IML (Lead) |
| Software | BS (Support), IML (Lead), CMF (Support) |
| Supervision | IML (Lead) |
| Validation | BS (Equal), CMF (Equal), KAB (Equal), RT (Equal), IML (Equal) |
| Visualization | BS (Support), IML (Lead) |
| Writing (Original Draft) | BS (Lead), IML (Support), RT (Support) |
| Writing (Review & Editing) | BS (Equal), CMF (Equal), JFC (Equal), KAB (Equal), RT (Equal), DN (Equal), IML (Equal) |

## Data Availability

All data and analysis code are available at: https://osf.io/4c8mb/ and data we extracted from the individual trials can be accessed here: https://bit.ly/3qFkcro

https://bit.ly/3qFkcro

## Footnotes

1 ^1^ According to the clinicaltrials.gov ‘history of changes’, the INTERVAL GAP-4 trial was registered on 31^st^ March 2016 with a trial start date of December 2015 and the trial status was ‘recruiting’. Although the trial start date was later changed on the 7^th^ September 2017 (for reasons unknown) to January 2016, this new date still means the trial was retrospectively preregistered by at least 12 weeks.

