## Supplementary tables and figures for "Outcome Reporting bias in Exercise Oncology trials (OREO): a cross-sectional study"

**OREO Supplementary tables and figures**

Supplementary Table 1. Search strategies

| PubMed search strategy for ClinicalTrials.gov, ISRCTN, EU Clinical Trials Register, ANZCTR, UMIN-CTR, NTR and ChiCTR registries (Search,Query, Items found) |
|---|
| #64,Search (#57 OR #58 OR #59 OR #60 OR #61 OR #62 OR #63),670<br>#63,Search (#47 AND #56),1<br>#62,Search (#47 AND #55),3<br>#61,Search (#47 AND #54),18<br>#60,Search (#47 AND #53),5<br>#59,Search (#47 AND #52),71<br>#58,Search (#47 AND #51),34<br>#57,Search (#47 AND #50),545<br>#56,Search UMIN CTR[Secondary Source ID],249<br>#55,Search ChiCTR[Secondary Source ID],686<br>#54,Search NTR[Secondary Source ID],1084<br>#53,Search EudraCT[Secondary Source ID],955<br>#52,Search ISRCTN[Secondary Source ID],7973<br>#51,Search ANZCTR[Secondary Source ID],2013<br>#50,Search clinicaltrials.gov[Secondary Source ID],76535<br>#47,Search (#43 NOT #46),9591<br>#46,Search (#44 NOT #45),4625192<br>#45,Search humans[MeSH Terms],18019805<br>#44,Search animals[MeSH Terms],22644997<br>#43,Search (#9 AND #33 AND #42),11071<br>#42,Search (#34 OR #35 OR #36 OR #37 OR #38 OR #39 OR #40 OR #41),4601787<br>#41,Search groups[Title/Abstract],1988377<br>#40,Search trial[Title/Abstract],565571<br>#39,Search randomly[Title/Abstract],319863<br>#38,Search drug therapy[MeSH Subheading],2145039<br>#37,Search placebo[Text Word],207732<br>#36,Search randomized[Title/Abstract],493662<br>#35,Search controlled clinical trial[Publication Type],579973<br>#34,Search randomized controlled trial[Publication Type],491521<br>#33,Search (#10 OR #11 OR #12 OR #13 OR #14 OR #15 OR #16 OR #17 OR #18 OR #19 OR #20 OR #21 OR #22 OR #23 OR #24 OR #25 OR #26 OR #27 OR #28 OR #29 OR #30 OR #31 OR #32),732813<br>#32,Search dancing[Title/Abstract],1694<br>#31,Search dance*[Title/Abstract],5378<br>#30,Search pilates[Title/Abstract],445<br>#29,Search qigong[Title/Abstract],665<br>#28,Search tai chi[Title/Abstract],1521<br>#27,Search yoga[Title/Abstract],4382<br>#26,Search running[MeSH Terms],19286<br>#25,Search cycling[Title/Abstract],57621<br>#24,Search bicycling[MeSH Terms],10725<br>#23,Search walk*[Title/Abstract],111057<br>#22,Search physical activities[Title/Abstract],6086<br>#21,Search physical activity[Title/Abstract],101752<br>#20,Search resistance training[MeSH Terms],7747<br>#19,Search sport*[Title/Abstract],73022<br>#18,Search Sports[Title/Abstract],47734<br>#17,Search motor activity[MeSH Terms],277023 |

#16,Search Exercis*[Title/Abstract],285554
#15,Search Muscle Strength[MeSH Terms],31129
#14,Search Physical Endurance[MeSH Terms],32286
#13,Search Physical Fitness[MeSH Terms],28082
#12,Search Exercise Movement Techniques[MeSH Terms],7724
#11,Search Exercise therapy[MeSH Terms],47483
#10,Search Exercise[MeSH Terms],183470
#9,Search (#1 OR #2 OR #3 OR #4 OR #5 OR #6 OR #7 OR #8),4142714
#8,Search tumour*[Title/Abstract],262250
#7,Search tumor*[Title/Abstract],1385999
#6,Search cancer*[Title/Abstract],1689341
#5,Search malignan*[Title/Abstract],548996
#4,Search adenocarcinoma*[Title/Abstract],141138
#3,Search carcinoma*[Title/Abstract],640787
#2,Search neoplas*[Title/Abstract],386933
#1,Search neoplasms[MeSH Terms],3223634

Registry search strategy for the WHO ICTRP registry.

Search 1
Search terms: Cancer AND Exercise
Phases: All
Results: All

Search 2
Search terms: Cancer AND physical activity
Phases: All
Results: All

* Exercise and physical activity terms were searched for separately

Supplementary Table 2. OREO Decisions during date entry (summary of decisions that required author discussions)

---

1) EXCITE trial (NCT01186367):

- Outcome: Exercise knowledge and exercise behaviour
  - **Decision: "Publication outcome analysis score matches registration? = No";** Because Exercise knowledge and exercise behaviour were pre-registered separately, but reported as a combined outcome
- Exercise behaviour was pre-registered as using the theory of planned behaviour therefore self-reported Physical activity using the Godwin was considered a new/novel outcome.
- Health status European Quality of Life-Five Dimension (EQ-5D) [Time Frame: Baseline, post intervention (2 months post baseline), 2 and 4 months follow up (post intervention completion)]:
  - **Decision: "Outcome score pre-registered?= Yes";** because this reflects the overall score from the questionnaire (i.e. not any sub-scale or domain scores)

---

2) Bridging the gap trial (NCT03087461)

- Exercise knowledge and exercise behaviour were pre-registered as separate outcomes but reported in the publication as one (combined) outcome. Therefore, exercise knowledge was considered correctly reported in publications (*with a note that it was combined with exercise behaviour) and exercise behaviour was considered missing.

---

3) DISPO trial (NCT01409720)

- Fatigue is assessed using the EORTC FA13 questionnaire [Time Frame: 12- and 24-weeks post completion of therapy]:
- **Decision: "Outcome score pre-registered? = No";** because this survey has multiple sub-scales and they have not specified the scales/score in the pre-registration.

---

4) PACT trial (NTR2138)

- Health related quality of life EORTC QOL C30, EQ5D at 0, 18w and 9mo; was listed was listed in the registry as one outcome, but is 2 separate outcomes (therefore entered separately as 2 outcomes)
- EORTC QOL C30
  - **Decision: "Outcome score pre-registered? = Yes";** because this represents the overall (total) QOL score. Therefore, only the overall score is considered pre-registered and all subscales/domains are considered as novel (non-reregistered)
- EQ5D
  - **Decision: "Outcome score pre-registered? = No";** because it is not specified whether it is the overall score, or a specific domain.
- Physical fitness (aerobic peak capacity, muscle strength): was listed in the registry as one outcome, but is 2 separate outcomes (therefore entered separately as 2 outcomes)

---

5) Loudon et al (ACTRN12611000202965)

- Quality of Life Questionnaire - LYMQOL:
  - **Decision: "Outcome score pre-registered? = No";** because Total QoL score +

---

domains reported in publication but "Quality of life questionnaire" pre-registered (did not specify whether it was overall score or specific domains), therefore all outcomes related to QOL were considered novel outcomes.

6) Newton et al (ACTRN12610000691044)

- Quality of life will be assessed using the EORTC QLQ-C30 was pre-registered. However, QLQ-PR25 (all subscales) were reported in the publication. Therefore, these were considered novel because the specific subscales of the QLQ-PR25 were not in the pre-registration
- HRQL (SF-36) was also reported in the publication. These were not in the pre-registration therefore listed as novel (non-preregistered) outcomes.
- Body composition (regional and whole-body fat mass and lean mass): registered as one outcome but represents separate outcomes.

7) ENGAGE trial (ACTRN12610000609055)

- Cancer-specific QOL EORTCQLQ-C30 V3 and Prostate tumour-specific module (EORTCQLQ-PR25) are 2 surveys, therefore were entered as 2 separate outcomes.
    - **Decision (for both): "Outcome score pre-registered?= Yes";** because this survey has an overall score (overall score was pre-registered). Any additional subscales

8) Winters-Stone et al (NCT00954044)

- Physical performance battery was reported in the paper as a composite score. However, **selected** individual components of the battery were also reported as separate individual outcomes (strength, chair stand and gait speed). Therefore, strength, chair stand and gait speed were also treated as non-preregistered outcomes. Note: the battery composite score included balance, however balance was not reported as a separate outcome.

9) SUPPORT trial (NCT01977066)

- Overall quality of life EORTC QLQ-C30; Change from 0, 6m:
    - **Decision: "Outcome score pre-registered? = Yes";** because this survey has an overall score. Therefore, the overall score was considered pre-registered, and any sub-scales were considered not pre-registered .
- Disease specific quality of life for pancreatic cancer (QLQ-PAN26);
    - **Decision: "Outcome score pre-registered? = Yes";** because this survey has an overall score. Therefore, the overall score was considered pre-registered, and any sub-scales were considered not pre-registered .
- **NOTE: Quality of life outcomes were not reported in publications. However, it is a recent trial (2019) and it is likely these are not yet published (will likely be published in the near future).

10) Newton et al (ACTRN12612000097842)

- Volumetric BMD and Bone Architecture are two separate outcomes
- Static and dynamic balance will be assessed using the Neurocom Smart Balancemaster. Baseline, 6 months and 12 months:
    - **Decision: "Measurement method pre-registered? = No":** because specific

method/protocol not reported (Neurocom Smart Balancemaster can provide various outcomes)

11) REACT trial (NL2036/NTR2153):

- Body composition; body height, body weight, waist and hip circumferences and four skinfolds (biceps, triceps, suprailiac and subscapular) at baseline, 12 weeks, and 1-year follow-up: listed as one outcome in the registry but it's 3 separate outcomes (1. body weight; 2. waist and hip circumference; 3. sum of four skinfolds)
- Functioning in daily life assessed using the Participation and Autonomy (IPA) Questionnaire at baseline, 12 weeks, and 1-year follow-up
    - **Decision: "Outcome score pre-registered? = No";** because cannot determine whether it is an overall (total) score, or specific domains. In the registry they report that the Participation and Autonomy (IPA) Questionnaire will be used. However, cannot determine which measure of physical functioning they will use from this questionnaire.
    - They registered Functioning in daily life, however Physical functioning was reported in publication (therefore, = Domain not reported).
- Mood disturbance; Hospital Anxiety and Depression Scale (HADS) at baseline, 12 weeks, and 1-year follow-up
    - **Decision: "Outcome score pre-registered?= No";** because cannot determine which mood disturbance outcome
    - No Mood disturbance outcomes reported in paper: therefore, it is considered as not reported
- Anxiety and depression were considered non pre-registered outcomes because they are deemed a different outcome to mood disturbance.
- Blood pressure: If a trial reported just blood pressure in the registration, we did not consider this as preregistering an outcome score. We required systolic, diastolic, or mean arterial blood pressure to be explicitly named.
- VO2 max: A trial was required to stipulate relative or absolute in name or provide units (l/min or ml/kg/min) that clarified which measure of VO2 max was used as the outcome score.

Supplementary Table 3. Completeness of statistical analysis results reporting

**Method:**

Finally, for each outcome that was reported in an eligible published manuscript, we noted the completeness of the reporting of statistical analysis for primary and secondary outcomes. We defined a result as completely reported if it was described in the publication to a level that the results could be used in a meta-analysis.

**Results:**

Almost all (98.8%) of the statistical analysis results of prespecified outcomes with an outcome score in published manuscripts were reported completely (i.e. reported to a level that the results could be used in a meta-analysis). All analyses of primary outcomes prespecified with an outcome score provided in published articles were reported completely. Researchers partially reported the analysis of five preregistered secondary outcomes with outcome scores (i.e. with only *p*-values) and failed to report just one (due to too few patients completing pre- and post-intervention assessments). All analysis results of insufficiently preregistered or novel outcomes were reported completely in published manuscripts, apart from five (0.5%) partially reported outcome analyses.

Supplementary Table 4. List of eligible trials (with registration and completion dates)

| Study ID | Primary investigator | Trial acronym | Registration no. | CR Ref | Date of registration dd/mm/yyyy | Study start date dd/mm/yyyy | Study completion date dd/mm/yyyy | Published protocol reference (n=18) | N cross-sectional papers (n=10) | N eligible results papers (n=78) | Publication references |
|---|---|---|---|---|---|---|---|---|---|---|---|
| 1 | Mustian | EXCAP | NCT00924651 | (1) | 19/6/2009 | 1/9/2009 | 1/10/2016 | None | 0 | 3 | (2–4) |
| 2 | Dieli-Conwright | None | NCT02454777 | (5) | 27/5/2015 | 29/9/2015 | 29/9/2021 | (6) | 0 | 4 | (7–10) |
| 3 | Keicolt-Glaser | None | NCT00486525 | (11) | 14/6/2007 | 1/8/2007 | 1/3/2013 | None | 0 | 2 | (12,13) |
| 4 | Galvão | None | ACTRN12611001158954 | (14) | 4/11/2011 | 28/08/2012 | NR | (15) | 1 | 1 | (16) |
| 5 | Littman | None | NCT00476203 | (17) | 21/5/52007 | 1/5/2007 | 1/5/2011 | None | 0 | 1 | (18) |
| 6 | Bridevaux | LCRS | NCT01258478 | (19) | 13/12/2010 | 1/12/2010 | 1/1/2014 | None | 0 | 3 | (20–22) |
| 7 | Schmitz | WISER | NCT01515124 | (23) | 23/1/2012 | 1/1/2012 | 1/5/2016 | (24) | 1 | 1 | (25) |
| 8 | Schmitz | PAL | NCT00194363 | (26) | 19/9/2005 | 1/10/2005 | 1/7/2007 | (27) | 1 | 6 | (28–33) |
| 9 | Cramer | YoCo | NCT01669109 | (34) | 20/8/2012 | 1/9/2012 | 1/12/2013 | None | 0 | 1 | (35) |
| 10 | Bjerre | Prostate FC | NCT02430792 | (36) | 30/4/2015 | 1/5/2015 | 1/2/2018 | (37) | 0 | 2 | (38,39) |
| 11 | Courneya | HITTS | NCT02459132 | (40) | 1/6/2015 | 1/7/2015 | 1/4/2017 | None | 0 | 2 | (41,42) |
| 12 | Furzer | THRIVING | ACTRN12609000450213 | (43) | 12/06/2009 | 1/08/2009 | NR | None | 0 | 1 | (44) |
| 13 | Dieli-Conwright | None | NCT01140282 | (45) | 9/6/2010 | 21/5/2012 | 1/10/2018 | (46) | 0 | 4 | (47–50) |
| 14 | Courneya | CARE | NCT00249015 | (51) | 7/11/2005 | 1/1/2008 | 1/3/2014 | None | 1 | 5 | (52–56) |
| 15 | Scott | EXCITE | NCT01186367 | (57) | 23/8/2010 | 1/8/2010 | 1/1/2021 | (58) | 0 | 1 | (59) |

| | | | | | | | | | | | |
|---|---|---|---|---|---|---|---|---|---|---|---|
| 16 | Smith-Turchyn | Bridging the Gap | NCT03087461 | (60) | 22/3/2017 | 1/6/2017 | 1/10/2018 | (61) | 0 | 2 | (62,63) |
| 17 | Rogers | BEAT | NCT00929617 | (64) | 29/6/2009 | 1/6/2009 | 1/12/2020 | (65) | 2 | 4 | (66–69) |
| 18 | Newton | None | ACTRN12610000691044 | (70) | 20/08/2010 | 27/06/2011 | 30/10/2012 | None | 0 | 1 | (71) |
| 19 | Rydwik | None | NCT02895464 | (72) | 9/9/2016 | 1/9/2016 | 30/6/2018 | None | 0 | 1 | (73) |
| 20 | Rogers | ABLE | NCT01147367 | (74) | 22/6/2010 | 1/8/2010 | 1/12/2013 | None | 0 | 3 | (75–77) |
| 21 | Debus | DISPO | NCT01409720 | (78) | 4/8/2011 | 1/9/2011 | 1/9/2013 | (79) | 0 | 7 | (80–86) |
| 22 | Velthuis | PACT | NTR2138 | (87) | 09/12/2009 | 01/01/2010 | 31/12/2012 | (88) | 1 | 4 | (89–92) |
| 23 | Loudon | None | ACTRN12611000202965 | (93) | 21/02/2011 | 22/02/2011 | NR | (94) | 0 | 2 | (95,96) |
| 24 | Livingston | ENGAGE | ACTRN12610000609055 | (97) | 27/07/2010 | 5/10/2011 | 28/06/2013 | (98) | 1 | 4 | (99–102) |
| 25 | Winters-Stone | None | NCT00954044 | (103) | 6/8/2009 | 1/1/2010 | 1/6/2011 | (104) | 0 | 1 | (105) |
| 26 | Greil | AGMT | NCT01384838 | (106) | 29/6/2011 | 1/6/2011 | 1/4/2015 | None | 0 | 1 | (107) |
| 27 | Steindorf | SUPPORT | NCT01977066 | (108) | 6/11/2013 | 1/11/2013 | 1/12/2016 | None | 0 | 2 | (109,110) |
| 28 | van Waart | PACES | NTR2159 | (111) | 11/01/2010 | 01/02/2010 | 31/07/2012 | (112) | 1 | 3 | (113–115) |
| 29 | O’Donnell | PEACH | NCT01030887 | (116) | 14/12/2009 | 1/1/2010 | 1/8/2011 | (117) | 0 | 2 | (118,119) |
| 30 | Newton | None | ACTRN12612000097842 | (120) | 20/01/2012 | 1/06/2012 | NR | (121) | 0 | 1 | (122) |
| 31 | Buffart | REACT | NTR2153 (NL2036) | (123) | 05/01/2010 | 01/03/2010 | 01/03/2014 | (124) | 1 | 3 | (125–127) |

Supplementary Table 5. List of full-text exclusions with reason for exclusion (n=143)

| | **Principal Investigator or first author** | **Trial acronym** | **Trial registration no.** | **Reason for exclusion** |
|---|---|---|---|---|
| 1 | DeAlmeida | None | NCT01693172 | Retrospectively registered |
| 2 | Mustian | YOCAS | NCT00397930 | Retrospectively registered |
| 3 | Wengstrom | OptiTrain | NCT02522260 | Retrospectively registered |
| 4 | Alibhai | ADT Ex RCT | NCT02046837 | Retrospectively registered |
| 5 | Steindorf | BEST | NCT01468766 | Retrospectively registered |
| 6 | Irwin | HOPE | NCT02056067 | Retrospectively registered |
| 7 | Rørth | None | NCT01711892 | Retrospectively registered |
| 8 | Courneya | HELP | NCT00111865 | Retrospectively registered |
| 9 | Courneya | START | NCT00115713 | Retrospectively registered |
| 10 | Gungorduk | None | NCT03553121 | Retrospectively registered |
| 11 | Belka | None | NCT03524755 | Retrospectively registered |
| 12 | Li | None | NCT02754973 | Retrospectively registered |
| 13 | Garcia | None | NCT03061773 | Retrospectively registered |
| 14 | Jones | None | NCT01725633 | Retrospectively registered |
| 15 | Schroeder | None | NCT01909440 | Retrospectively registered |
| 16 | Finn | ExPeCT | NCT02453139 | Retrospectively registered |
| 17 | Knobf | None | NCT01102985 | Retrospectively registered |

| | | | | |
|---|---|---|---|---|
| 18 | Grüllich | None | NCT01645150 | Retrospectively registered |
| 19 | Campbell | None | NCT01296893 | Retrospectively registered |
| 20 | Villanueva | None | NCT02052050 | Retrospectively registered |
| 21 | Dunne | None | NCT01523353 | Retrospectively registered |
| 22 | Santa Mina | None | NCT02036684 | Retrospectively registered |
| 23 | Edvardsen | FALC | NCT01748981 | Retrospectively registered |
| 24 | Winters-Stone | None | NCT00660686 | Retrospectively registered |
| 25 | Pedersen | PROLUCA | NCT01893580 | Retrospectively registered |
| 26 | Karvinen | None | NCT01130714 | Retrospectively registered |
| 27 | Jones | None | NCT00620932 | Retrospectively registered |
| 28 | Quist | EXHALE | NCT01881906 | Retrospectively registered |
| 29 | Jarden | PACE-AL | NCT01404520 | Retrospectively registered |
| 30 | Carli | None | NCT00227526 | Retrospectively registered |
| 31 | LaStayo | None | NCT00335491 | Retrospectively registered |
| 32 | Yeo | None | NCT00902759 | Retrospectively registered |
| 33 | Oldervoll | None | NCT00397774 | Retrospectively registered |
| 34 | Moadel | None | NCT00179348 | Retrospectively registered |
| 35 | Thomas | POSITIVE | NCT02055508 | Retrospectively registered |
| 36 | Lai | None | ChiCTR-IOR-16008109 | Retrospectively registered |

| | | | | |
|---|---|---|---|---|
| 37 | Christensen | PROTRACT | ISRCTN32132990 | Retrospectively registered |
| 38 | Arbane | None | ISRCTN/ISRCTN07216922 | Retrospectively registered |
| 39 | Adamsen | None | ISRCTN05322922 | Retrospectively registered |
| 40 | Mutrie | None | ISRCTN12587864 | Retrospectively registered |
| 41 | Galvao | None | ACTRN12609000200280 | Retrospectively registered |
| 42 | Newton | INTERVAL-GAP4 | NCT02730338 | Retrospectively registered |
| 43 | Bender | EPICC | NCT02793921 | Retrospectively registered |
| 44 | Weid | SURfit | NCT02730767 | Retrospectively registered |
| 45 | Meneveau | CARDAPAC | NCT02433067 | Retrospectively registered |
| 46 | Bohus | PETRA | NCT01374399 | Retrospectively registered |
| 47 | Courneya | ERASE | NCT03203460 | No published results (published protocol only) |
| 48 | Allen | None | NCT02950324 | No published results (published protocol only) |
| 49 | Mikkelsen | PACE-Mobil-PBL | NCT03411200 | No published results (published protocol only) |
| 50 | McIsaac | PREHAB | NCT02934230 | No published results (published protocol only) |
| 51 | FILAIRE | Rexochir | NCT03020251 | No published results (published protocol only) |
| 52 | Jack | EMPOWER | NCT01914068 | No published results (published protocol only) |
| 53 | Cuesta-Vargas | EFICATEST | NCT02433197 | No published results (published protocol only) |
| 54 | Hammel | APACaP | NCT02184663 | No published results (published protocol only) |
| 55 | Liu | ANITA | ACTRN12617000975392 | No published results (published protocol only) |
| 56 | Bruce | PROSPER | ISRCTN35358984 | No published results (published protocol only) |

| | | | | |
|---|---|---|---|---|
| 57 | | | | No published results |
| 58 | Carayol | APAD | NCT01495650 | Includes nutrition intervention |
| 59 | Foucaut | PASAPAS | NCT01331772 | Includes nutrition intervention |
| 60 | Schink | None | NCT02293239 | Includes nutrition intervention |
| 61 | Bourke | None | ISRCTN88605738 | Includes nutrition intervention |
| 62 | van Gemert | SHAPE-2 | NCT01511276 | Includes nutrition intervention |
| 63 | Haseen | None | ISCRTN75282423 | Includes nutrition intervention |
| 64 | Cohen | None | NCT03171506 | Includes nutrition intervention |
| 65 | Focht | IDEA-P | NCT02050906 | Includes nutrition intervention |
| 66 | Yun | LEACH | NCT01527409 | Includes nutrition intervention |
| 67 | Daley | None | ISRCTN/ISRCTN08045231 | Includes nutrition intervention |
| 68 | Kiechle | LIBRE-1 | NCT02087592 | Includes nutrition intervention |
| 69 | Arikawa | None | NCT02940470 | Includes nutrition intervention |
| 70 | van der Werf | None | NTR4223 | Includes nutrition intervention |
| 71 | Stacey | ENRICH | ANZCTRN1260901086257 | Includes nutrition intervention |
| 72 | Eriksen | None | NCT01300104 | Includes nutrition intervention |
| 73 | Frugé | None | NCT01886677 | Includes nutrition intervention |
| 74 | Saxton | None | ISRCTN08045231 | Includes nutrition intervention |
| 75 | Le Roy | PREHAB | NCT02780921 | Includes nutrition intervention |

| | | | | |
|---|---|---|---|---|
| 76 | Tsuruta | None | NCT02224807 | Includes nutrition intervention |
| 77 | Thomson | LIVES | NCT00719303 | Includes nutrition intervention |
| 78 | Kapoor | None | NCT02350855 | Includes nutrition intervention |
| 79 | Demark-Wahnefried | None | NCT01886677 | Includes nutrition intervention |
| 80 | Ho | None | NCT01708824 | Includes nutrition intervention |
| 81 | Schink | None | NCT02293239 | Includes nutrition intervention |
| 82 | Kenzik | None | NCT01112839 | Includes nutrition intervention |
| 83 | Gnagnarella | InForma | NCT02622711 | Includes nutrition intervention |
| 84 | Solheim | MENAC | NCT02330926 | Includes nutrition intervention |
| 85 | Pan | None | NCT03268629 | Non-exercise intervention |
| 86 | Bewarder | None | NCT03467087 | Non-exercise intervention |
| 87 | Bolli | SENECA | NCT02509156 | Non-exercise intervention |
| 88 | Jibb | None | NCT02611739 | Non-exercise intervention |
| 89 | Litt | CAPs | NCT03089177 | Non-exercise intervention |
| 90 | Huhn | None | NCT02621554 | Non-exercise intervention |
| 91 | McDermott | None | NCT01176487 | Non-exercise intervention |
| 92 | Esplen | ReBIC | NCT00418444 | Non-exercise intervention |
| 93 | Rades | (PRE-MODE | NCT03070431 | Non-exercise intervention |
| 94 | Tambour | None | NCT02015897 | Non-exercise intervention |

| | | | | |
|---|---|---|---|---|
| 95 | Zernicke | eCALM | NCT01476891 | Non-exercise intervention |
| 96 | Arving | None | NCT01588262 | Non-exercise intervention |
| 97 | Andersen | None | NCT00990977 | Non-exercise intervention |
| 98 | Würtzen | None | NCT00990977 | Non-exercise intervention |
| 99 | Baker | None | ACTRN12611001094965 | Non-exercise intervention |
| 100 | Sherman | None | ACTRN12615001381572 | Non-exercise intervention |
| 101 | Maddocks | None | ISRCTN42944026 | Non-exercise intervention |
| 102 | Dowswell | None | ISRCTN03320951 | Non-exercise intervention |
| 103 | Challand | None | ISRCTN14680495 | Non-exercise intervention |
| 104 | Torres Lacomba | None | ISRCTN95870846 | Non-exercise intervention |
| 105 | Streckmann | VANISH | NCT03032718 | Non-exercise intervention |
| 106 | Trinh | RiseTx | NCT03321149 | Distance-based (web-based intervention) |
| 107 | Ritvo | iMOVE | NCT02620735 | Distance-based (web-based intervention) |
| 108 | Pfirrmann | iPEP | NCT02478996 | Distance-based (web-based intervention) |
| 109 | Valle | FITNET | NCT01349153 | Distance-based (web-based intervention) |
| 110 | Fazzino | None | NCT01441011 | Distance-based (web-based intervention) |
| 111 | Fu | None | NCT02462226 | Distance-based (web-based intervention) |
| 112 | Fergus | None | NCT01089764 | Distance-based (web-based intervention) |
| 113 | Lee | None | NCT01512069 | Distance-based (web-based intervention) |

| | | | | |
|---|---|---|---|---|
| 114 | Huang | Fit4Life | NCT01253720 | Distance-based (web-based intervention) |
| 115 | Quintiliani | None | NCT02387671 | Distance-based (web-based intervention) |
| 116 | Bantum | None | NCT00962494 | Distance-based (web-based intervention) |
| 117 | Valle | None | NCT01349153 | Distance-based (web-based intervention) |
| 118 | Golsteijn | None | NTR4296 | Distance-based (web-based intervention) |
| 119 | Kanera | None | NTR3375 | Distance-based (web-based intervention) |
| 120 | Glynn | SMART MOVE | ISRCTN/ISRCTN99944116 | Distance-based (web-based intervention) |
| 121 | Ormel | SMART-trial | NCT02391454 | Distance-based (web-based intervention) |
| 122 | Paxton | None | NCT02722850 | Distance-based (web-based intervention) |
| 123 | Thorsen | None | NCT01749774 | Distance-based/ Behavioural based (no face to face exercise supervision) |
| 124 | McNeil | None | NCT03564899 | Distance-based |
| 125 | Vallance | None | NCT00221221 | Distance-based |
| 126 | Vallerand | None | NCT03052777 | Distance-based |
| 127 | Hartman | None | NCT02332876 | Distance-based |
| 128 | Pinto | InForma | NCT0262271, ISRCTN53325751 | Distance-based |
| 129 | Gordon | None | ACTRN12608000399392 | Distance-based |
| 130 | Van Blarigan | Smart Pace | NCT02966054 | Distance-based |
| 131 | Dittus | None | NCT01482702 | Behavioural based |
| 132 | Prinsen | None | NCT01096641 | Behavioural based |

| | | | | |
|---|---|---|---|---|
| 133 | Procter | None | ISRCTN72882329 | Behavioural based |
| 134 | Gielissen | None | NCT01096641 | Behavioural based |
| 135 | Hathiramani | REIL | NCT02272751 | No face-to-face supervision |
| 136 | Winters-Stone | None | NCT03120819 | No face-to-face supervision |
| 137 | Gokal | None | ISRCTN50709297 | No face-to-face supervision |
| 138 | Morey | RENEW | NCT00303875 | No face-to-face supervision |
| 139 | Lahart | None | NCT02408107 | No face-to-face supervision |
| 140 | Moug | REx trial | ISRCTN62859294 | No face-to-face supervision |
| 141 | Winters-Stone | None | NCT03120819 | No face-to-face supervision |
| 142 | Cheema | None | ANZCTR/12612000346875 | Wrong outcome |
| 143 | Weiner | None | NCT02332876 | Wrong outcome |

Supplementary Table 6. Trials that were retrospectively registered (but otherwise eligible for inclusion) (n=46)

| | Principal Investigator | Trial acronym | Trial registration no. | Date of registration | Trial start date[1] | N days retrospectively registered | Publications |
|---|---|---|---|---|---|---|---|
| 1 | DeAlmeida | None | NCT01693172 | September 26, 2012 | August 2012[2] | 56 | Early mobilization programme improves functional capacity after major abdominal cancer surgery: a randomized controlled trial. |
| 2 | Mustian | YOCAS | NCT00397930 | November 10, 2006 | October 2006 | 40 | Influence of Yoga on Cancer-Related Fatigue and on Mediational Relationships Between Changes in Sleep and Cancer-Related Fatigue: A Nationwide, Multicenter Randomized Controlled Trial of Yoga in Cancer Survivors.<br><br>Multicenter, randomized controlled trial of yoga for sleep quality among cancer survivors. |
| 3 | Wengstrom | OptiTrain | NCT02522260 | August 13, 2015 | March 2013 | 895 | Two-year follow-up of the OptiTrain randomised controlled exercise trial.<br><br>Adding high-intensity interval training to conventional training modalities: optimizing health-related outcomes during chemotherapy for breast cancer: the OptiTrain randomized controlled trial.<br><br>Optitrain: a randomised controlled exercise trial for women with breast cancer undergoing chemotherapy. |
| 4 | Alibhai | ADT Ex RCT | NCT02046837 | January 28, 2014 | November 2013 | 88 | A phase II randomized controlled trial of three exercise delivery methods in men with prostate cancer on androgen deprivation therapy.<br><br>A phase II RCT and economic analysis of three exercise delivery methods in men with prostate cancer on androgen deprivation therapy. |

| | | | | | | | |
|---|---|---|---|---|---|---|---|
| 5 | Steindorf | BEST | NCT01468766 | November 9, 2011 | February 2011 | 281 | Effects of exercise on sleep problems in breast cancer patients receiving radiotherapy: a randomized clinical trial.<br><br>Factors influencing participation in a randomized controlled resistance exercise intervention study in breast cancer patients during radiotherapy.<br>Randomized, controlled trial of resistance training in breast cancer patients receiving adjuvant radiotherapy: results on cancer-related fatigue and quality of life.<br><br>Randomized controlled trial to evaluate the effects of progressive resistance training compared to progressive muscle relaxation in breast cancer patients undergoing adjuvant radiotherapy: the BEST study.<br>Resistance Exercise Reduces Kynurenine Pathway Metabolites in Breast Cancer Patients Undergoing Radiotherapy |
| 6 | Irwin | HOPE | NCT02056067 | February 5, 2014 | June 2009 | 1710 | The effect of exercise on body composition and bone mineral density in breast cancer survivors taking aromatase inhibitors.<br>Randomized exercise trial of aromatase inhibitor-induced arthralgia in breast cancer survivors.<br><br>Randomized exercise trial of aromatase inhibitor-induced arthralgia in breast cancer survivors. |
| 7 | Rørth | None | NCT01711892 | October 22, 2012 | March 2012 | 235 | Football training in men with prostate cancer undergoing androgen deprivation therapy: activity profile and short-term skeletal and postural balance adaptations.<br><br>Effects of recreational soccer in men with prostate cancer undergoing androgen deprivation therapy: study protocol for the 'FC Prostate' randomized controlled trial. |

| | | | | | | | |
|---|---|---|---|---|---|---|---|
| | | | | | | | Efficacy of recreational football on bone health, body composition, and physical functioning in men with prostate cancer undergoing androgen deprivation therapy: 32-week follow-up of the FC prostate randomised controlled trial.<br><br>Structural and functional cardiac adaptations to a 10-week school-based football intervention for 9-10-year-old children.<br><br>"All boys and men can play football": a qualitative investigation of recreational football in prostate cancer patients.<br><br>Football training improves lean body mass in men with prostate cancer undergoing androgen deprivation therapy. |
| 8 | Courneya | HELP | NCT00111865 | May 27, 2005 | April 2005 | 56 | Patient satisfaction with participation in a randomized exercise trial: effects of randomization and a usual care posttrial exercise program.<br><br>A randomized trial of aerobic exercise and sleep quality in lymphoma patients receiving chemotherapy or no treatments.<br><br>Effects of supervised exercise on motivational outcomes and longer-term behavior. |
| 9 | Courneya | START | NCT00115713 | June 24, 2005 | April 2003 | 815 | Effects of exercise during adjuvant chemotherapy on breast cancer outcomes.<br><br>Hemoglobin and aerobic fitness changes with supervised exercise training in breast cancer patients receiving chemotherapy.<br><br>Understanding breast cancer patients' preference for two |

| | | | | | | | |
|---|---|---|---|---|---|---|---|
| | | | | | | | types of exercise training during chemotherapy in an unblinded randomized controlled trial. |
| 10 | Gungorduk | None | NCT03553121 | June 12, 2018 | January 1, 2018 | 162 | Impact of pre-operative walking on post-operative bowel function in patients with gynecologic cancer. |
| 11 | Belka | None | NCT03524755 | May 15, 2018 | July 10, 2013 | 1770 | Progressive resistance training in cachectic head and neck cancer patients undergoing radiotherapy: a randomized controlled pilot feasibility trial. |
| 12 | Li | None | NCT02754973 | April 28, 2016 | May 2015 | 363 | An integrated experiential training programme with coaching to promote physical activity, and reduce fatigue among children with cancer: A randomised controlled trial. |
| 13 | Garcia | None | NCT03061773 | February 23, 2017 | March 1, 2014 | 1090 | Effect of exercise on pain and functional capacity in breast cancer patients. |
| 14 | Jones | None | NCT01725633 | November 14, 2012 | December 2010 | 714 | Feasibility, safety, and efficacy of aerobic training in pretreated patients with metastatic breast cancer: A randomized controlled trial. |
| 15 | Schroeder | None | NCT01909440 | July 26, 2013 | July 8, 2013 | 18 | Impact of resistance training on body composition and metabolic syndrome variables during androgen deprivation therapy for prostate cancer: a pilot randomized controlled trial.<br><br>A pilot randomised controlled trial of a periodised resistance training and protein supplementation intervention in prostate cancer survivors on androgen deprivation therapy. |
| 16 | Finn | ExPeCT | NCT02453139 | May 25, 2015 | October 2014 | 236 | The ExPeCT (Examining Exercise, Prostate Cancer and Circulating Tumour Cells) trial: study protocol for a randomised controlled trial.<br><br>The views of patients with metastatic prostate cancer towards physical activity: a qualitative exploration. |

| | | | | | | | |
|---|---|---|---|---|---|---|---|
| 17 | Knobf | None | NCT01102985 | April 13, 2010 | January 2008 | 833 | The Yale Fitness Intervention Trial in female cancer survivors: Cardiovascular and physiological outcomes. |
| 18 | Grüllich | None | NCT01645150 | July 20, 2012 | May 2012 | 80 | Resistance training as supportive measure in advanced cancer patients undergoing TKI therapy-a controlled feasibility trial. |
| 19 | Campbell | None | NCT01296893 | February 16, 2011 | January 2011 | 46 | Effect of aerobic exercise on cancer-associated cognitive impairment: A proof-of-concept RCT. |
| 20 | Villanueva (Morales) | None | NCT02052050 | January 31, 2014 | September 2012 | 517 | Effectiveness of Lumbopelvic Exercise in Colon Cancer Survivors: A Randomized Controlled Clinical Trial. |
| 21 | Dunne | None | NCT01523353 | February 1, 2012 | July 2011 | 215 | Randomized clinical trial of prehabilitation before planned liver resection. |
| 22 | Santa Mina | None | NCT02036684 | January 15, 2014 | November 2013 | 75 | Prehabilitation for men undergoing radical prostatectomy: a multi-centre, pilot randomized controlled trial.<br><br>Prehabilitation and acute postoperative physical activity in patients undergoing radical prostatectomy: a secondary analysis from an RCT. |
| 23 | Edvardsen | FALC | NCT01748981 | December 13, 2012 | November 2010 | 773 | High-intensity training following lung cancer surgery: a randomised controlled trial.<br><br>Reduction in cardiorespiratory fitness after lung resection is not related to the number of lung segments removed. |
| 24 | Winters-Stone | None | NCT00660686 | April 17, 2008 | January 2006 | 837 | Resistance training reduces disability in prostate cancer survivors on androgen deprivation therapy: evidence from a randomized controlled trial. |
| 25 | Pedersen | PROLUCA | NCT01893580 | July 9, 2013 | May 2012 | 434 | Perioperative rehabilitation in operation for lung cancer (PROLUCA) - rationale and design. |
| 26 | Karvinen | None | NCT01130714 | May 26, 2010 | January | 145 | Effect of an exercise training intervention with |

| | | | | | | | |
|---|---|---|---|---|---|---|---|
| | | | | | 2010 | | resistance bands on blood cell counts during chemotherapy for lung cancer: a pilot randomized controlled trial. |
| 27 | Jones | None | NCT00620932 | February 22, 2008 | January 2008 | 52 | Effects of nonlinear aerobic training on erectile dysfunction and cardiovascular function following radical prostatectomy for clinically localized prostate cancer. |
| 28 | Quist | EXHALE | NCT01881906 | June 20, 2013 | February 2012 | 505 | "EXHALE": exercise as a strategy for rehabilitation in advanced stage lung cancer patients: a randomized clinical trial comparing the effects of 12 weeks supervised exercise intervention versus usual care for advanced stage lung cancer patients.<br><br>Effects of an exercise intervention for patients with advanced inoperable lung cancer undergoing chemotherapy: A randomized clinical trial.<br><br>Early initiated postoperative rehabilitation enhances quality of life inpatients with operable lung cancer: Secondary outcomes from a randomized trial. |
| 29 | Jarden | PACE-AL | NCT01404520 | July 28, 2011 | June 2011 | 57 | Patient Activation through Counseling and Exercise--Acute Leukemia (PACE-AL)--a randomized controlled trial.<br><br>Multimodal intervention integrated into the clinical management of acute leukemia improves physical function and quality of life during consolidation chemotherapy: a randomized trial 'PACE-AL'. |
| 30 | Carli | None | NCT00227526 | September 28, 2005 | February 2005 | 239 | Impact of preoperative change in physical function on postoperative recovery: argument supporting prehabilitation for colorectal surgery.<br><br>Randomized clinical trial of prehabilitation in colorectal surgery. |

| | | | | | | | |
|---|---|---|---|---|---|---|---|
| 31 | LaStayo | None | NCT00335491 | June 12, 2006 | March 2006 | 103 | Eccentric exercise versus usual-care with older cancer survivors: the impact on muscle and mobility--an exploratory pilot study. |
| 32 | Yeo | None | NCT00902759 | May 15, 2009 | January 9, 2009 | 126 | A progressive postresection walking program significantly improves fatigue and health-related quality of life in pancreas and periampullary cancer patients. |
| 33 | Oldervoll | None | NCT00397774 | November 10, 2006 | October 2006 | 40 | Physical exercise for cancer patients with advanced disease: a randomized controlled trial. |
| 34 | Moadel | None | NCT00179348 | September 16, 2005 | February 8, 2001 | 1681 | Randomized controlled trial of yoga among a multiethnic sample of breast cancer patients: effects on quality of life. |
| 35 | Thomas | POSITIVE | NCT02055508 | February 5, 2014 | December 2013 | 66 | POSITIVE study: physical exercise program in non-operable lung cancer patients undergoing palliative treatment. |
| 36 | Lai | None | ChiCTR-IOR-16008109 | March 16, 2016 | January 2015 | 440 | Systematic short-term pulmonary rehabilitation before lung cancer lobectomy: a randomized trial. |
| 37 | Christensen | PROTRACT | ISRCTN32132990 | March 8, 2011 | January 2011 | 66 | Safety and efficacy of resistance training in germ cell cancer patients undergoing chemotherapy: a randomized controlled trial. |
| | | | | | | | Progressive resistance training and cancer testis (PROTRACT) - efficacy of resistance training on muscle function, morphology and inflammatory profile in testicular cancer patients undergoing chemotherapy: design of a randomized controlled trial. |
| 38 | Arbane | None | ISRCTN/ISRCTN07216922 | September 21, 2010 | September 2010 | 20 | Effect of postoperative physical training on activity after curative surgery for non-small cell lung cancer: a multicentre randomised controlled trial. |
| 39 | Adamsen | None | ISRCTN05322922 | July 5, 2007 | October 2003 | 1373 | Effect of a multimodal high intensity exercise intervention in cancer patients undergoing chemotherapy: randomised controlled trial |

| | | | | | | | |
|---|---|---|---|---|---|---|---|
| 40 | Mutrie | None | ISRCTN12587864 | September 26, 2005 | January 2004 | 628 | Benefits of supervised group exercise programme for women being treated for early stage breast cancer: pragmatic randomised controlled trial. |
| 41 | Galvao | None | ACTRN12609000200280 | April 21, 2009 | October 2008 | 202 | Exercise Improves VO2max and Body Composition in Androgen Deprivation Therapy-treated Prostate Cancer Patients. |
| 42 | Newton | INTERVAL-GAP4 | NCT02730338 | April 6, 2016 | December 2015 | 127 | Intense Exercise for Survival among Men with Metastatic Castrate-Resistant Prostate Cancer (INTERVAL-GAP4): a multicentre, randomised, controlled phase III study protocol |
| 43 | Bender | EPICC | NCT02793921 | June 7, 2016 | April 2016 | 67 | Protocol for Exercise Program in Cancer and Cognition (EPICC): A Randomized Controlled Trial of the Effects of Aerobic Exercise on Cognitive Function in Postmenopausal Women With Breast Cancer Receiving Aromatase Inhibitor Therapy<br><br>Physical activity, cardiorespiratory fitness, and cognitive function in postmenopausal women with breast cancer |
| 44 | Von Der Weid | SURfit | NCT02730767 | March 31, 2016 | August 2015 | 243 | SURfit - A Physical Activity Intervention for Childhood Cancer Survivors (SURfit) |
| 45 | Meneveau | CARDAPAC | NCT02433067 | May 04, 2015 | April 2015 | 33 | Physical Activity Intervention on Myocardial Function in Patients With HER2 + Breast Cancer (CARDAPAC) |
| 46 | Bohus | PETRA | NCT01374399 | June 16, 2011 | February 2011 | 135 | Physical Exercise Therapy vs Relaxation in Allogeneic Stem Cell Transplantation (PETRA) |

[1] The 1st of the month was used as the date if only the month (without a date) was reported in the registry.

[2] Recruited for almost 2 and a half years (August 2012 to 11th January 2015), before the authors moved the start date from August 2012 to December 2014, and made extensive changes to the exercise intervention, outcome measures, and eligibility criteria thereafter. In the period between changes, on four separate occasions, the authors made changes to the trial outcomes and design, but not the trial recruitment status or the trial start date. We, therefore, rejected the change in trial start date. However, because the registration was first submitted (September 2012) before the original start date (August 2012), the trial was retrospectively registered.

Supplementary Table 7. Trials with no published results but were otherwise eligible for inclusion (n=11)

| # | Author (PI) | Acronym | Registration number | Date of registration | Trial start date | Study completion date | Days since or until completion (As of January 22, 2021) | Trial Status | Link to published protocol | Link to registration |
|---|---|---|---|---|---|---|---|---|---|---|
| 1 | Courneya | ERASE | NCT03203460 | June 29, 2017 | July 1, 2018 | May 31, 2021 | -129 | Ongoing | https://www.ncbi.nlm.nih.gov/pubmed/31278095 | https://clinicaltrials.gov/ct2/show/NCT03203460 |
| 2 | Allen | None | NCT02950324 | November 1, 2016 | November 2016 | November 2022 | -648 | Ongoing | https://www.ncbi.nlm.nih.gov/pubmed/30580268 | https://clinicaltrials.gov/ct2/show/NCT02950324 |
| 3 | Mikkelsen | PACE-Mobil-PBL | NCT03411200 | January 26, 2018 | April 4, 2018 | July 2020 | 205 | Completed | https://www.ncbi.nlm.nih.gov/pubmed/30261853 | https://clinicaltrials.gov/ct2/show/NCT03411200 |
| 4 | McIsaac | PREHAB | NCT02934230 | October 14, 2016 | January 5, 2017 | March 2021 | -38 | Ongoing | https://www.ncbi.nlm.nih.gov/pubmed/29934394 | https://clinicaltrials.gov/ct2/show/NCT02934230 |
| 5 | FILAIRE | Rexochir | NCT03020251 | January 13, 2017 | May 12, 2017 | September 2021 | -222 | Ongoing | https://www.ncbi.nlm.nih.gov/pubmed/29133320 | https://clinicaltrials.gov/ct2/show/NCT03020251 |
| 6 | Jack | EMPOWER | NCT01914068 | August 1, 2013 | August 2013 | December 2015 | 1879 | Completed | https://www.ncbi.nlm.nih.gov/pubmed/26762365 | https://clinicaltrials.gov/ct2/show/NCT01914068 |
| 7 | Cuesta-Vargas | EFICATEST | NCT02433197 | May 4, 2015 | December 2015 | December 2019 | NA | Suspended | https://www.ncbi.nlm.nih.gov/pubmed/267 | https://clinicaltrials.gov/ct2/show/NCT024 |

| | | | | | | | | | | |
|---|---|---|---|---|---|---|---|---|---|---|
| | | | | | | | | | 32120 | 33197 |
| 8 | Hammel | APACaP | NCT02184663 | July 9, 2014 | October 15, 2014 | December 2021 | -313 | Ongoing | https://www.ncbi.nlm.nih.gov/pubmed/26458923 | https://clinicaltrials.gov/ct2/show/NCT02184663 |
| 9 | Liu | ANITA | ACTRN12617000975392 | July 6, 2017 | August 1, 2017 | February 2019 | 721 | Ongoing | https://www.ncbi.nlm.nih.gov/pubmed/29526243 | https://anzctr.org.au/Trial/Registration/TrialReview.aspx?ACTRN=ACTRN12617000975392 |
| 10 | Bruce | PROSPER | ISRCTN35358984 | February 4, 2015 | March 1, 2015 | March 2020 | 314 | Completed | https://www.ncbi.nlm.nih.gov/pubmed/29574439 | http://www.isrctn.com/ISRCTN35358984 |
| 11 | Hayes | SAFE | ACTRN12616000547448 | April 28, 2016 | May 2, 2016 | February 2, 2018 | 1085 | Completed | None | http://anzctr.org.au/Trial/Registration/TrialReview.aspx?ACTRN=12616000547448 |

Supplementary Table 8. Characteristics of included trials

| | Overall | Clinical trials.gov | ANZ trials registry | Netherlands trial registry |
|---|---|---|---|---|
| N prospectively registered trials | 31 (100%) | 22 (71%) | 6 (19%) | 3 (10%) |
| Total publications[1] | | | | |
| • N total publications | 106 (100%) | 74 (70%) | 16 (15%) | 16 (15%) |
| • Median (min-max) publications per trial | 3 (1-8) | 3 (1-8) | 3 (1-6) | 5 (5-6) |
| Published protocols: | | | | |
| • Published protocol (yes) | 18 (100%) | 11 (61%) | 4 (22%) | 3 (17%) |
| • Prospectively published (yes) | 3 (100%) | 1 (33%) | 2 (67%) | 0 (0%) |
| • Retrospective published (yes) | 15 (100%) | 10 (67%) | 2 (13%) | 3 (20%) |
| Cross-sectional papers: | | | | |
| • N total publications | 10 (100%) | 5 (50%) | 2 (20%) | 3 (30%) |
| • Median (min-max) publications per trial | 0 (0-2) | 0 (0-2) | 0 (0-1) | 1 (1-1) |
| Results papers | | | | |
| • N total publications | 78 (100%) | 58 (74%) | 10 (13%) | 10 (13%) |
| • Median (min-max) publications per trial | 2 (1-7) | 2 (1-7) | 1 (1-4) | 3 (3-4) |
| Overall sample size: | | | | |
| • N participants across trials | 4,393 | 3,010 | 616 | 767 |
| • Mean (SD) | 137 ±116 | 137 ± 119 | 103 ± 110 | 192 ± 114 |
| • Median (min-max) | 65 (23-420) | 65 (23-420) | 60 (28-320) | 234 (23-277) |
| Exercise intervention groups: | | | | |
| • N participants | 2,393 | 1,626 | 293 | 473 |
| • Mean (SD) | 61 ± 49 | 63 ±52 | 49 ± 48 | 68 ± 43 |
| • Median (min-max) | 50 (7-231) | 43 (11-231) | 30 (15-142) | 77 (7-119) |
| Control groups: | | | | |
| • N participants | 1,950 | 1,333 | 323 | 294 |
| • Mean (SD) | 61 ± 53 | 61 ±54 | 54 ± 62 | 74 ± 47 |
| • Median (min-max) | 32 (8-225) | 32 (12-225) | 30 (13-178) | 84 (8-118) |
| N trials per cancer type: | | | | |
| • Breast | 13 (100%) | 12 (92%) | 1 (8%) | 0 (0%) |
| • Colorectal | 2 (100%) | 2 (100%) | 0 (0%) | 0 (0%) |
| • Breast and colorectal | 1 (100%) | 0 (0%) | 0 (0%) | 1 (100%) |
| • Haematological | 1 (100%) | 0 (0%) | 1 (100%) | 0 (0%) |
| • Lung | 1 (100%) | 1 (100%) | 0 (0%) | 0 (0%) |
| • Mixed[2] | 5 (100%) | 3 (60%) | 0 (0%) | 2 (40%) |
| • Pancreatic | 1 (100%) | 1 (100%) | 0 (0%) | 0 (0%) |
| • Prostate | 6 (100%) | 2 (33%) | 4 (66%) | 0 (0%) |
| • Testicular | 1 (100%) | 1 (100%) | 0 (0%) | 0 (0%) |
| Age, years: | | | | |
| • Mean ± SD | 58 ± 8 | 58 ± 8 | 63 ± 8 | 53 ± 4 |

| | | | | |
|---|---|---|---|---|
| • Median (Min-max) | 58 (44-76) | 57 (44-76) | 67 (49-70) | 52 (51-58) |
| Mean % of female patients per trial | 65 ± 41% | 73 ± 37% | 20 ± 45% | 83 ± 16% |
| Body mass index, kg/m$^2$ | | | | |
| • Mean ± SD | 28 ± 3 | 28 ± 3 | 28 ± 1 | 25 ± 1 |
| • Median (Min-max) | 28 (23-34) | 28 (23-34) | 28 (25-30) | 25 (24-26) |
| Ethnicity: | | | | |
| • Mean ± SD %, White per trial | 72 ± 26% | 72 ± 26% | NR | NR |
| • Mean ± SD %, Black per trial | 13 ± 12% | 13 ± 12% | NR | NR |
| • Mean ± SD %, Other per trial | 2 ± 2% | 2 ± 2% | NR | NR |
| • N (%) trials not reporting ethnicity | 18 (100%) | 9 (50%) | 6 (33%) | 3 (17%) |
| Cancer stage: | | | | |
| • Mean ± SD %, stage 0 | 4 ± 9% | 5 ± 11% | 1 ± 2% | 0 ± 0% |
| • Mean ± SD %, stage I | 26 ± 18% | 29 ± 17% | 23 ± 20% | 3 ± 4% |
| • Mean ± SD %, stage II | 33 ± 18% | 34 ± 17% | 27 ± 24% | 30 ± 24% |
| • Mean ± SD %, stage III | 24 ± 20% | 22 ± 17% | 11 ± 10% | 63 ± 22% |
| • Mean ± SD %, stage IV | 10 ± 28% | 7 ± 22% | 33 ± 58% | 5 ± 6% |
| • Mean ± SD %, unknown stage | 9 ± 23% | 11 ± 26% | 0 ± 0% | 0 ± 0% |
| • N trials not reporting stage | 5 (100%) | 1 (20%) | 3 (60%) | 1 (20%) |
| Treatment stage | | | | |
| • Pre-treatment (surgery) | 2 (100%) | 2 (100%) | 0 (0%) | 0 (0%) |
| • Undergoing chemotherapy, and/or radiotherapy | 9 (100%) | 6 (67%) | 1 (11%) | 2 (22%) |
| • Undergoing aromatase inhibitor treatment | 1 (100%) | 1 (100%) | 0 (0%) | 0 (0%) |
| • Undergoing ADT treatment | 2 (100%) | 0 (0%) | 2 (100%) | 0 (0%) |
| • Post-treatment (surgery, chemotherapy, and/or radiotherapy) | 17 (100%) | 13 (76%) | 3 (18%) | 1 (6%) |
| Intervention mode | | | | |
| • Aerobic exercise | 9 (100%)[3] | 9 (100%)[3] | 0 (0%) | 0 (0%) |
| • Resistance exercise | 5 (100%)[3] | 5 (100%)[3] | 0 (0%) | 0 (0%) |
| • Combined aerobic and resistance | 14 (100%) | 6 (43%) | 5 (36%) | 3 (21%) |
| • Yoga | 4 (100%) | 3 (75%) | 1 (25%) | 0 (0%) |
| Supervision | | | | |
| • Fully supervised | 26[4] (100%) | 18 (69%) | 5 (19%) | 3 (12%) |
| • Partly supervised[5] | 5 (100%) | 4 (80%) | 1 (20%) | 0 (0%) |
| Intervention duration (weeks) | | | | |
| • Mean ± SD | 18 ± 13 | 19 ± 15 | 14 ± 6 | 18 ± 6 |
| • Median (min-max) | 12 (3-52) | 14 (3-52) | 12 (8-26) | 18 (12-24) |

[1] Includes published protocols, cross-sectional papers and results papers.
[2] Mixed included mixed samples of participants diagnosed with breast, colorectal, oesophageal, gynaecological, lymphoma, cervix, testicular, lung, melanoma, renal, pancreatic, urogenital, adrenal or peritoneal cancer.
[3] One trial involved two exercise intervention arms (aerobic exercise intervention arm and a resistance exercise intervention arm).
[4] Three trials had an additional unsupervised intervention arm.
[5] Partly supervised was considered less than half of the exercise sessions involving in-person supervision.

Supplementary Table 9. Summary of outcomes published in trial results manuscripts

| | | N or % |
|---|---|---|
| General information: preregistered outcomes | Number of eligible trials | 31 |
| | Number of prespecified outcomes in registries and protocols | 405 |
| | Number of outcomes preregistered with an outcome score | 250 |
| | Proportion of outcomes preregistered with an outcome score | 61.7% |
| General information: published outcomes | Number of outcomes in published reports | 929 |
| | Number of outcomes preregistered with an outcome score that were published in trial reports | 158 |
| | Proportion of outcomes preregistered with an outcome score that were published in trial reports | 63.2% |
| | Proportion of outcomes in trial publications that were sufficiently preregistered outcomes | 17.0% |
| | Number of trials with no preregistered outcome score published in trial reports | 8 |
| | Proportion of trials with no preregistered outcome score published in trial reports | 25.8% |
| Primary outcomes | Number of preregistered primary outcomes in registries and protocols | 71 |
| | Proportion of preregistered outcomes in registries and protocols that were primary | 17.5% |
| | Median (min-max) primary outcomes per trial registry and protocol | 1 (1-8) |
| | Number of all outcomes labelled as primary across eligible trial publications | 170 |
| | Proportion of all outcomes across eligible trial publications that were labelled as primary | 18.3% |
| | Number of primary outcomes preregistered with an outcome score in registries and protocols | 39 |
| | Number of primary outcomes preregistered with an outcome score reported in published reports | 34 |
| | Proportion of primary outcomes preregistered with an outcome score in registries and protocols that were reported in published reports | 87.2% |
| | Proportion of all outcomes labelled as primary in published reports that were primary outcomes preregistered with an outcome score | 20.0% |
| Primary reported as primary | Number of primary outcomes preregistered with an outcome score correctly reported as primary outcomes in published reports | 28 |
| | Proportion of preregistered primary outcomes in trial registries and protocols reported as primary outcomes | 39.4% |
| | Proportion of all outcomes labelled as primary in trial reports that were primary outcomes preregistered with an outcome score | 16.5% |
| | Proportion of primary outcomes preregistered with an outcome score correctly reported as primary outcomes in published reports | 72% |
| | Proportion of all published outcomes that were primary outcomes preregistered with an outcome score reported as primary outcomes | 3.0% |

| | | |
|---|---|---|
| | Number of trials with no prespecified primary outcomes reported as a primary outcome in publications | 15 |
| | Proportion of trials with no prespecified primary outcomes reported as a primary outcome in publications | 48.4% |
| Primary reported as secondary | Number of primary outcomes preregistered with an outcome score reported as secondary outcomes in published reports | 6 |
| | Proportion of preregistered primary outcomes in trial registries and protocols reported as secondary outcomes | 8.5% |
| | Proportion of all outcomes labelled as secondary in trial reports that were primary outcomes preregistered with an outcome score | 0.8% |
| | Proportion of primary outcomes preregistered with an outcome score reported as secondary outcomes | 15.4% |
| | Proportion of all published outcomes that were primary outcomes preregistered with an outcome score reported as secondary outcomes | 0.6% |
| Unreported primary outcomes | Trials with no primary outcomes preregistered with an outcome score reported in trial publications | 13 |
| | Proportion of eligible trials that have no primary outcomes preregistered with an outcome score reported in trial publications | 41.9% |
| | Primary outcomes preregistered with an outcome score not reported in trial publications | 5 |
| | Proportion of primary outcomes preregistered with an outcome score in trial registries that were not reported in trial publications | 7.0% |
| Secondary Outcomes | Number of prespecified secondary outcomes in registries and protocols | 334 |
| | Proportion of prespecified outcomes in registries and protocols that were secondary | 82.5% |
| | Median (min-max) secondary outcomes per trial registry and protocol | 6 (0-58) |
| | Number of all outcomes labelled as secondary across eligible trial publications | 759 |
| | Proportion of all outcomes across eligible trial publications that were labelled as secondary | 81.7% |
| | Number of secondary outcomes preregistered with an outcome score in registries and protocols | 211 |
| | Number of secondary outcomes preregistered with an outcome score reported in published reports | 124 |
| | Proportion of secondary outcomes preregistered with an outcome score in registries and protocols that were reported in published reports | 58.8% |
| | Proportion of all outcomes labelled as secondary in published reports that were secondary outcomes preregistered with an outcome score | 16.3% |
| Secondary as secondary | Number of primary outcomes preregistered with an outcome score correctly reported as primary outcomes in published reports | 120 |
| | Proportion of preregistered primary outcomes in trial registries and protocols reported as primary outcomes | 35.9% |
| | Proportion of all outcomes labelled as primary in trial reports that were primary outcomes preregistered with an outcome score | 15.8% |
| | Proportion of primary outcomes preregistered with an outcome score correctly reported as primary outcomes in published reports | 56.9% |

| | | |
|---|---|---|
| | Proportion of all published outcomes that were primary outcomes preregistered with an outcome score reported as primary outcomes | 12.9% |
| | Number of trials with no prespecified primary outcomes reported as a primary outcome in publications | 10 |
| | Proportion of trials with no prespecified primary outcomes reported as a primary outcome in publications | 32.3% |
| Secondary reported as primary | Number of secondary outcomes preregistered with an outcome score reported as primary outcomes in published reports | 4 |
| | Proportion of preregistered secondary outcomes in trial registries and protocols reported as primary outcomes | 1.2% |
| | Proportion of all outcomes labelled as primary in trial reports that were secondary outcomes preregistered with an outcome score | 2.4% |
| | Proportion of secondary outcomes preregistered with an outcome score reported as primary outcomes | 1.9% |
| | Proportion of all published outcomes that were secondary outcomes preregistered with an outcome score reported as primary outcomes | 0.4% |
| Unreported secondary outcomes | Trials with no secondary outcomes preregistered with an outcome score reported in trial publications | 10 |
| | Proportion of eligible trials that have no secondary outcomes preregistered with an outcome score reported in trial publications | 32.3% |
| | Secondary outcomes preregistered with an outcome score not reported in trial publications | 87 |
| | Proportion of secondary outcomes preregistered with an outcome score in trial registries that were not reported in trial publications | 26.0% |
| Domain-only preregistered outcomes | Number of outcomes in registries and protocols that were domain-only preregistered outcomes | 155 |
| | Proportion of all outcomes in registries and protocols that were domain-only preregistered outcomes | 38.3% |
| | Median (min-max) domain-only preregistered outcomes per trial registry and protocol | 4 (0-18) |
| | Number of primary outcomes in registries and protocols that were domain-only preregistered outcomes | 32 |
| | Number of secondary outcomes in registries and protocols that were domain-only preregistered outcomes | 123 |
| | Number of outcomes in published reports that were related to domain-only preregistered outcomes | 328 |
| | Proportion of published outcomes that were related to domain-only preregistered outcomes | 35.3% |
| Primary domain-only outcomes in published reports | Number of primary outcomes in published reports that were related to domain-only preregistered outcomes | 75 |
| | Proportion of all outcomes in published reports that were primary outcomes related to domain-only preregistered outcomes | 8.1% |
| | Proportion of outcomes labelled as primary outcomes in published reports that were related to domain-only preregistered outcomes | 44.1% |
| Secondary domain-only outcomes in published reports | Number of secondary outcomes in published reports that were related to domain-only preregistered outcomes | 253 |

| | | |
|---|---|---|
| | Proportion of all outcomes in published reports that were secondary outcomes related to domain-only preregistered outcomes | 27.2% |
| | Proportion of outcomes labelled as secondary outcomes in published reports that were related to domain-only preregistered outcomes | 33.3% |
| Primary domain-only as primary | Number of primary outcomes preregistered as domain-only reported as primary outcomes in published reports | 55 |
| | Proportion of all published domain-only outcomes that were primary outcomes preregistered as domain-only reported as primary outcomes in published reports | 16.8% |
| | Proportion of all published outcomes that were primary outcomes preregistered as domain-only reported as primary outcomes in published reports | 5.9% |
| Primary domain-only as secondary | Number of primary outcomes preregistered as domain-only reported as secondary outcomes in published reports | 20 |
| | Proportion of all published domain-only that were primary outcomes preregistered as domain-only reported as secondary outcomes in published reports | 6.1% |
| | Proportion of all published outcomes that were primary outcomes preregistered as domain-only reported as secondary outcomes in published reports | 2.2% |
| Secondary domain-only as secondary | Number of secondary outcomes preregistered as domain-only reported as secondary outcomes in published reports | 197 |
| | Proportion of all published domain-only that were secondary outcomes preregistered as domain-only reported as secondary outcomes in published reports | 60.1% |
| | Proportion of all published outcomes that were secondary outcomes preregistered as domain-only reported as secondary outcomes in published reports | 21.2% |
| Secondary domain-only as primary | Number of secondary outcomes preregistered as domain-only reported as primary outcomes in published reports | 56 |
| | Proportion of all published domain-only that were secondary outcomes preregistered as domain-only reported as primary outcomes in published reports | 17.1% |
| | Proportion of all published outcomes that were secondary outcomes preregistered as domain-only reported as primary outcomes in published reports | 6.0% |
| Unreported domain-only preregistered outcomes | Number of unreported domain-only preregistered outcomes across eligible trials | 74 |
| | Proportion of all outcomes in registries and protocols that were unreported primary domain-only preregistered outcomes | 18.3% |
| | Median (min-max) unreported domain-only preregistered outcomes per trial | 1 (0-16) |
| | Number of unreported domain-only preregistered primary outcomes | 4 |
| | Proportion of primary outcomes in registries and protocols that were unreported primary domain-only preregistered outcomes | 5.6% |
| | Number of unreported domain-only preregistered secondary outcomes | 70 |
| | Proportion of all secondary outcomes in registries and protocols that were unreported secondary domain-only preregistered outcomes | 21.0% |

| | | |
|---|---|---|
| Non-preregistered (novel) Outcomes | Total number of novel outcomes | 443 |
| | Proportion of all published outcomes that were novel outcomes | 47.7% |
| | Median (min-max) novel outcomes, per trial | 9 (0-69) |
| | Number of trials with no novel outcomes | 4 |
| | Proportion of eligible trials with no novel outcomes | 12.9% |
| | Number of novel outcomes reported as primary outcomes in published reports | 63 |
| | Proportion of novel outcomes that were reported as primary outcomes in published reports | 14.2% |
| | Number of novel outcomes reported as secondary outcomes in published reports | 380 |
| | Proportion of novel outcomes that were reported as secondary outcomes in published reports | 85.8% |
| | Number of novel outcomes declared as novel | 49 |
| | Proportion of novel outcomes that were declared as novel | 11.1% |
| | Number of novel primary outcomes declared as novel | 3 |
| | Number of novel secondary outcomes declared as novel | 46 |
| | Number of trials with novel outcomes that were all declared | 1 |
| | Proportion of trials with novel outcomes that were all declared | 3.2% |
| | Median (min-max) novel outcomes reported without declaration per trial | 14 (0-50) |

Supplementary Table 10. Main deviations from our preregistered protocol

| Preregistered plan | Deviation from preregistered plan with rationale |
|---|---|
| Categorisation of the level of outcome registration and preregistered outcome scores | We introduced a process for:<br>1. the categorisation of the level of outcome registration as either "outcome domain only", "partial" or "complete".<br>2. the categorisation of pre-registered outcome scores as either "domain plus method", "outcome score" or "domain only"<br><br>We introduced this during the data-entry process in order to provide greater detail about the quality of outcome pre-registration and to allow us to identify the sources of selective outcome reporting and switching. This represents a necessary deviation from our pre-planned approach for the current study.<br><br>Our intended approach of including only outcomes with prespecified measurement methods, outcome scores, assessment timepoints, and descriptions of how the metrics were calculated and used, would have yielded too few outcomes for our analysis. In addition, this deviation allowed us to account for all outcomes reported in eligible trial publications. Therefore, we adopted the less conservative approach of distinguishing between outcomes with and without a prespecified outcome score. |
| If an outcome is prespecified at several timepoints, each timepoint will be considered as a separate outcome. | We did not consider each time point will be considered as a separate outcome, instead we noted whether assessments for each timepoint were reported at each outcome. |
| Data entry | Use of multiple authors for data entry (instead of a single author). |
| Article type | Exclusion of cross-sectional and mediation analysis publications from our main analyses. |
| Exploratory analysis | Odds of a domain-related outcome being switched versus an outcome preregistered with an outcome score. |

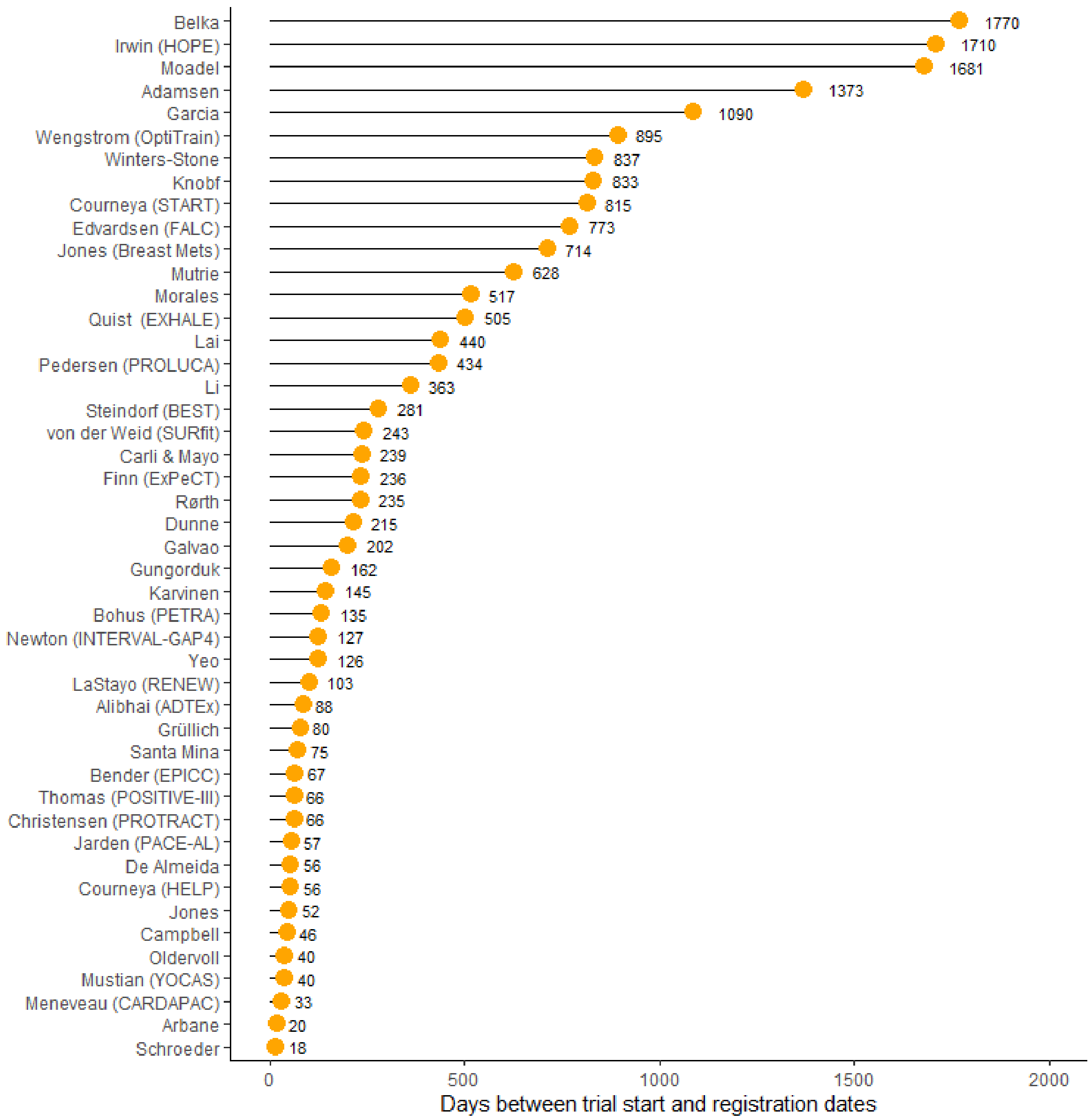


Supplementary figure 1. Days between trial start and registration for each retrospectively registered trial (trials are identified by the lead investigators name with trial acronym, if available). The PROLUCA and CARDAPAC trials were terminated, and Jones was withdrawn.

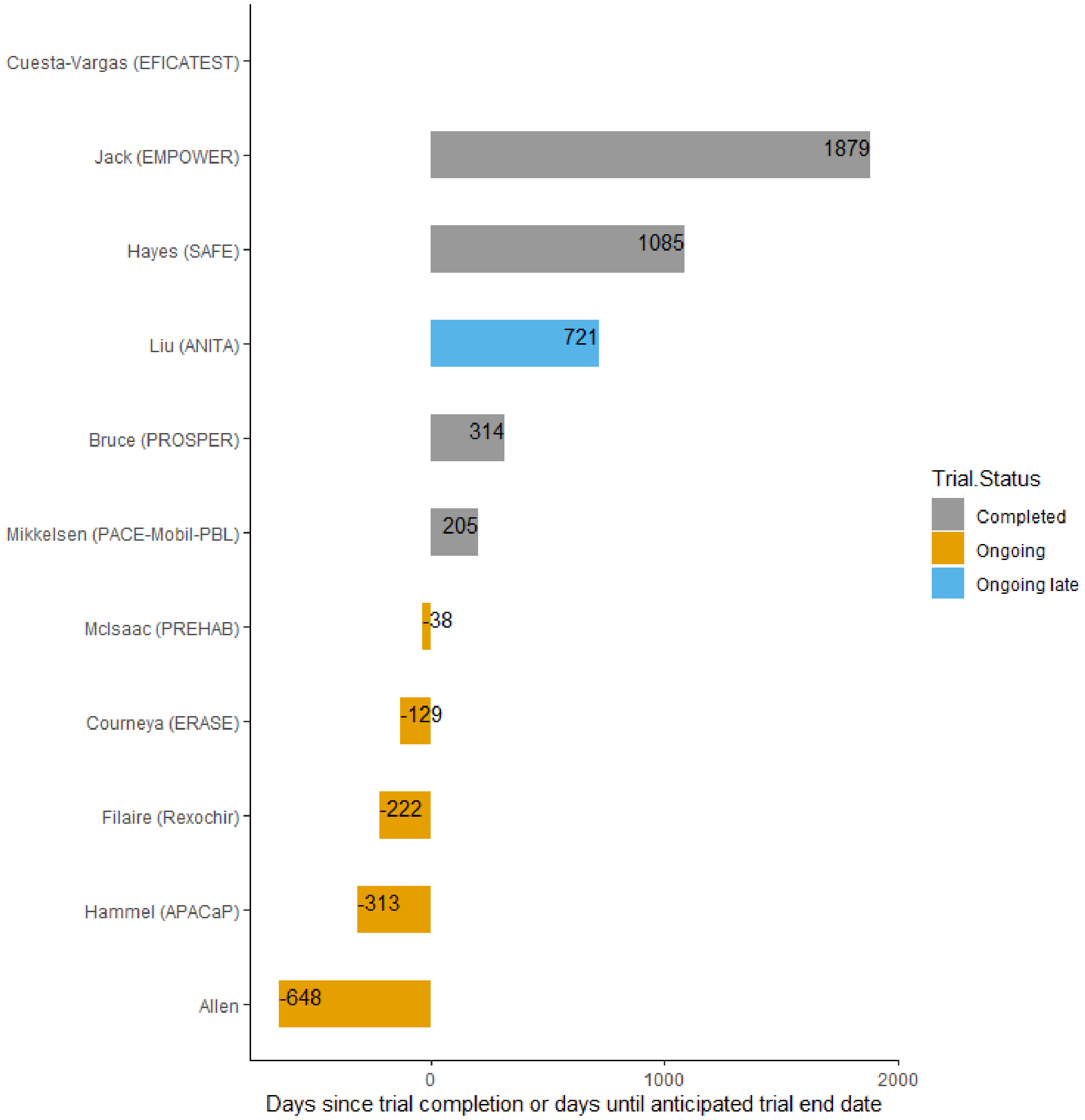


Supplementary figure 2. The number of days from trial completion until November 2020 for the completed trials (in red) and days from November 2020 until the anticipated trial completion date for ongoing trials (in green). The number of days since the anticipated trial completion date was plotted for the ANITA trial. No data are available for the EFICATEST trial because it was suspended.

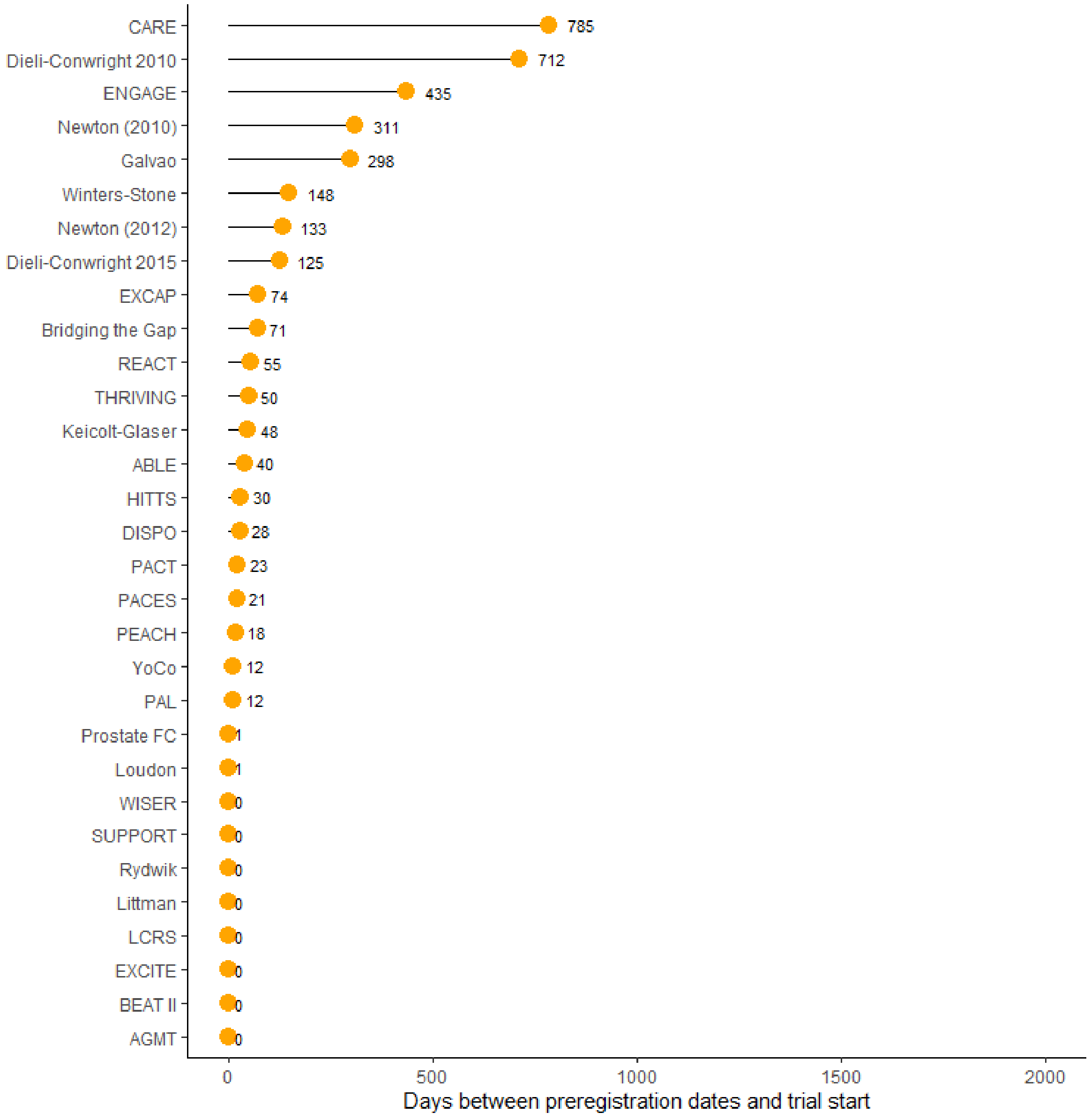


Supplementary figure 3. Days between preregistration dates and trial start

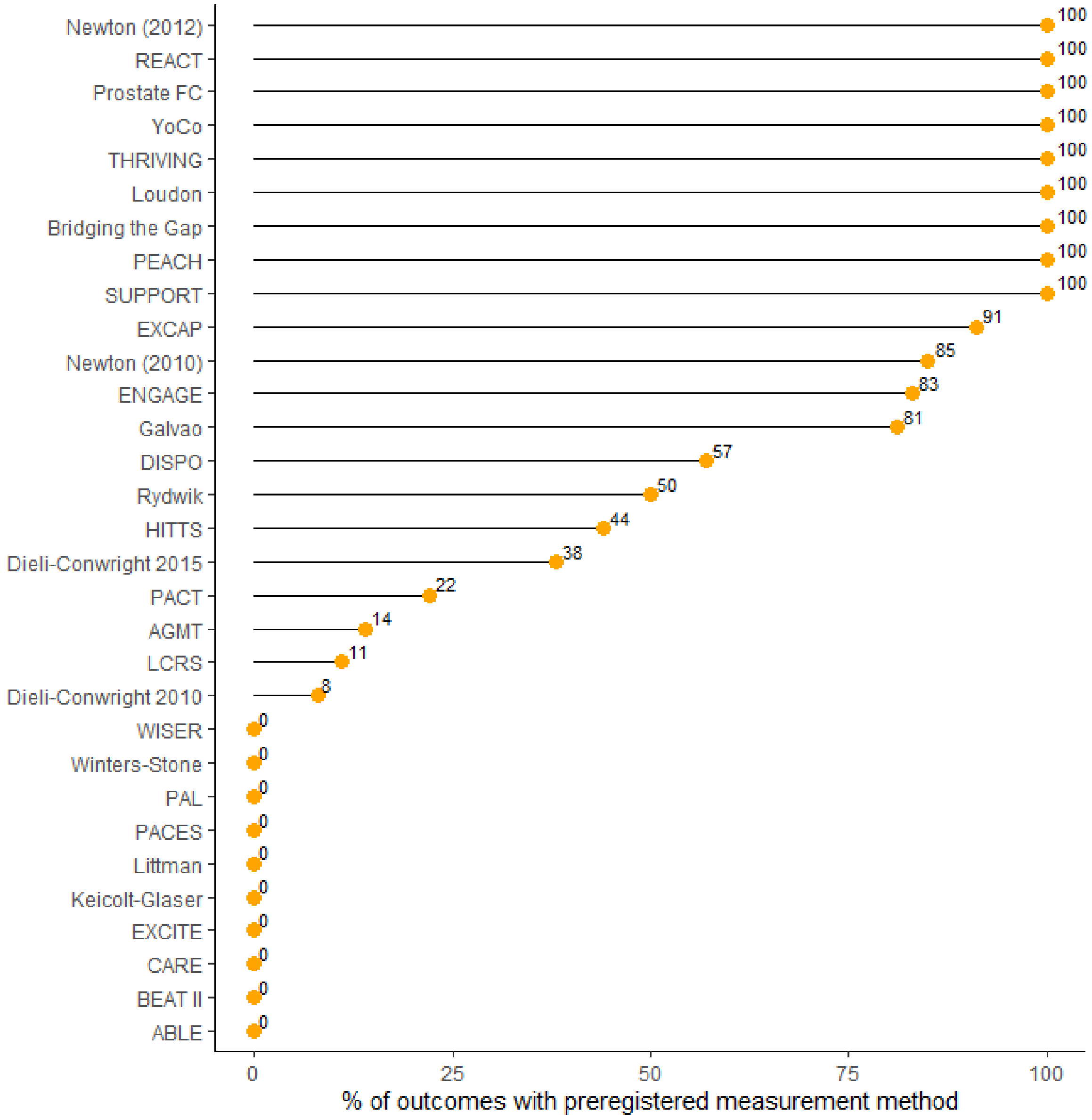


Supplementary figure 4. Percentage of outcomes preregistered with a measurement method

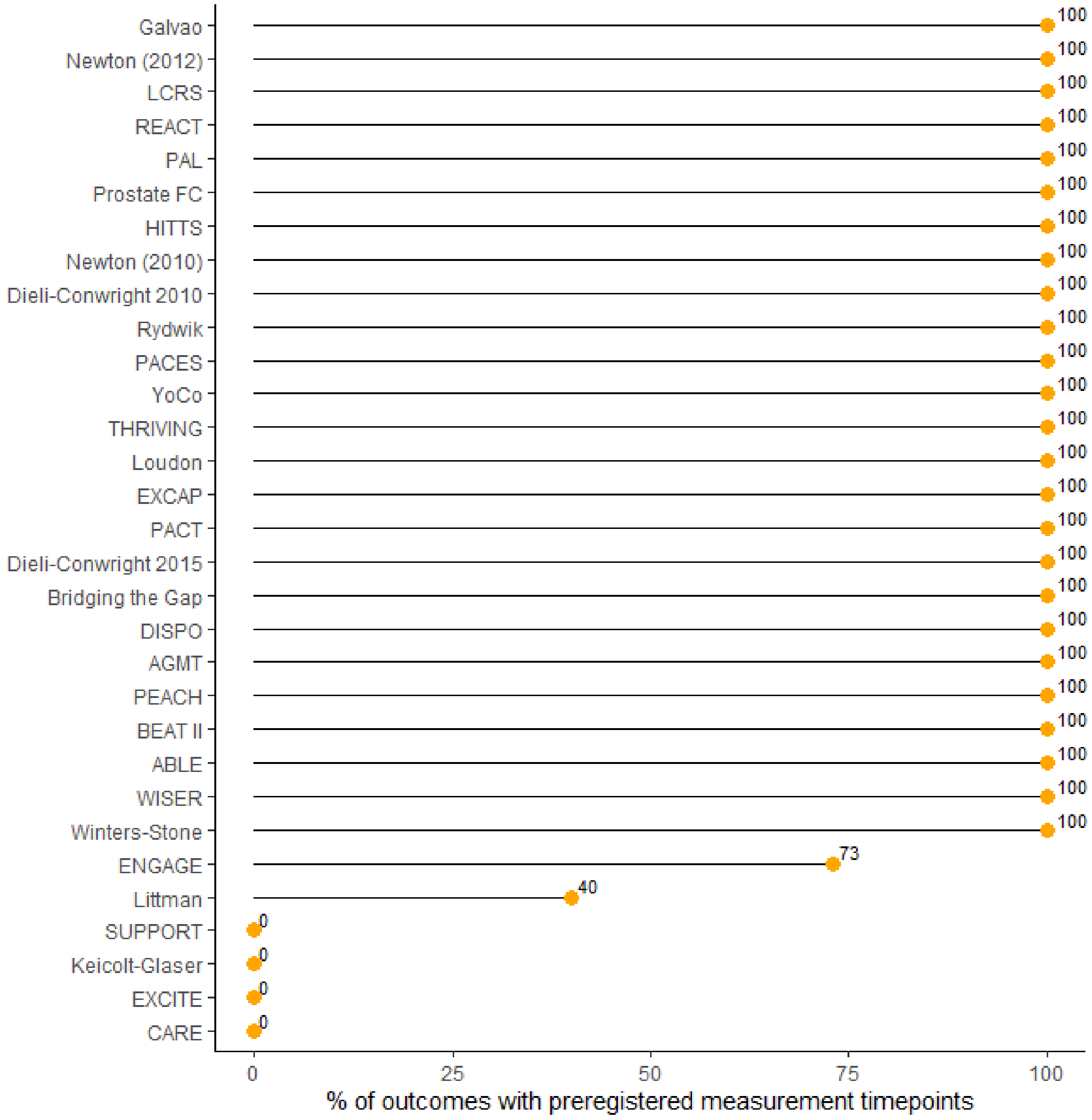


Supplementary figure 5. Percentage of outcomes preregistered with a measurement timepoints

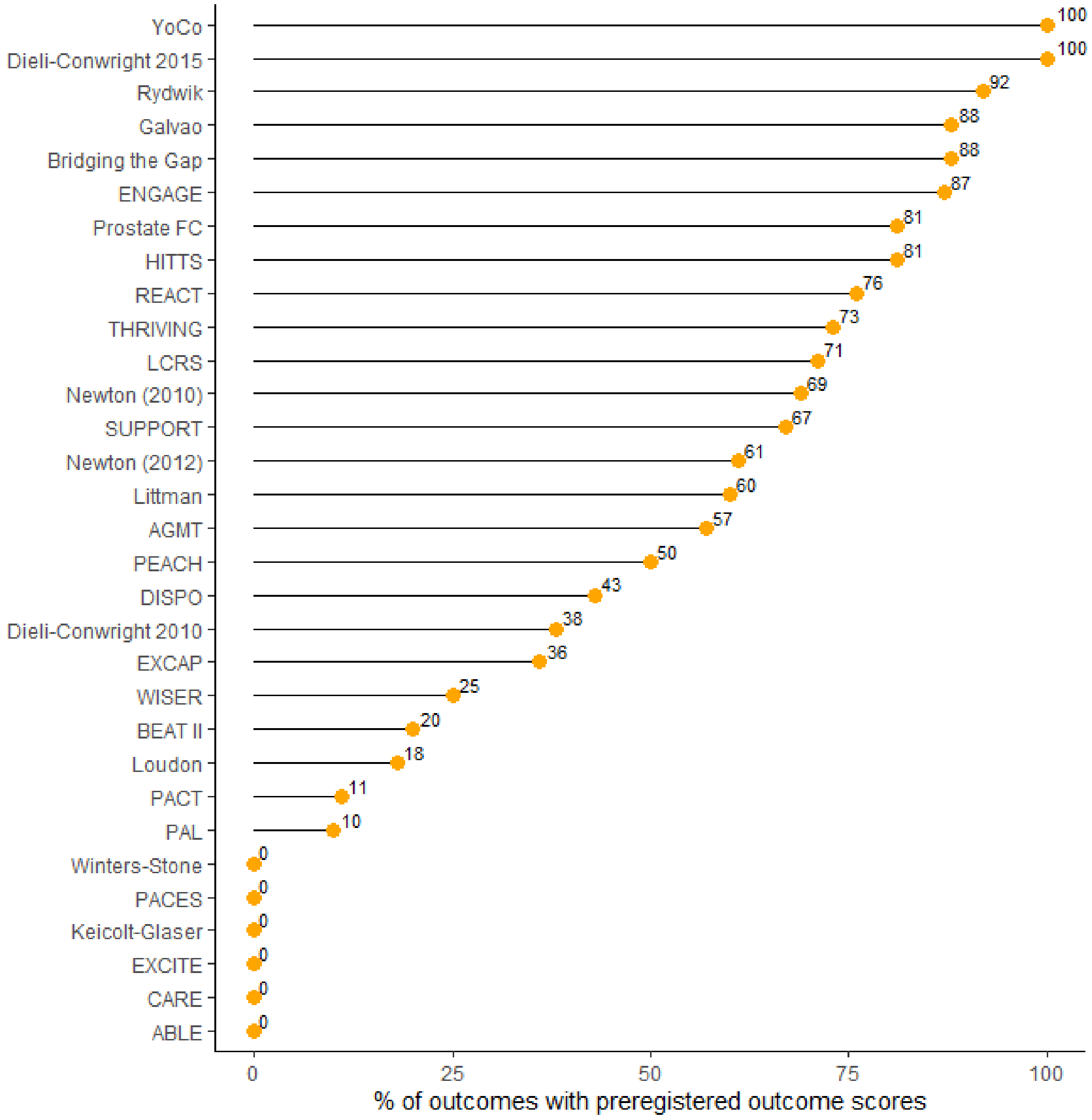


Supplementary figure 6. Percentage of outcomes preregistered with an outcome score

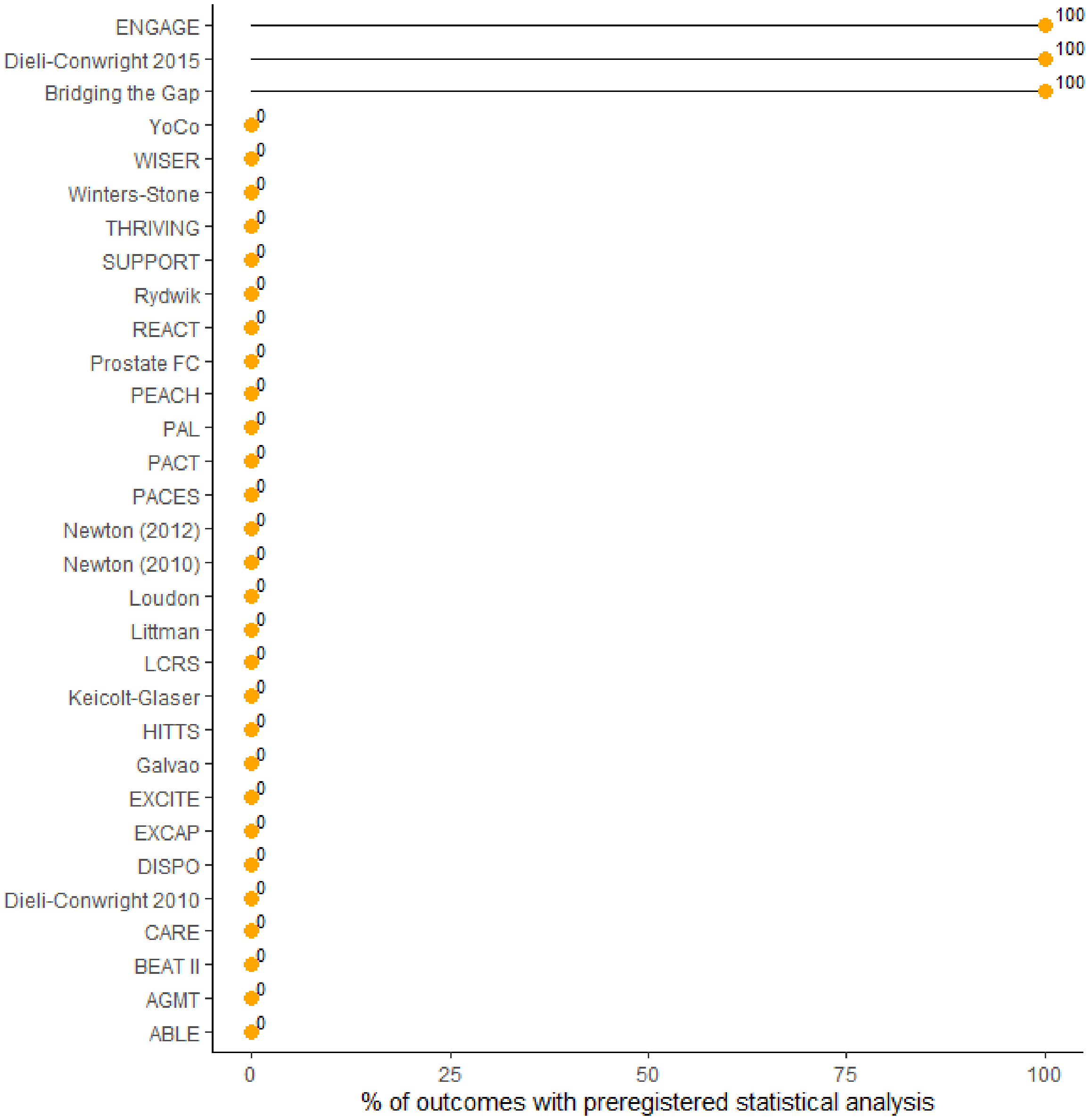


Supplementary figure 7. Percentage of outcomes preregistered with a statistical analysis description
